# Knowledge augmented causal discovery through large language models and knowledge graphs: application in chronic low back pain

**DOI:** 10.64898/2026.02.13.26346255

**Authors:** Damon Lin, Marzieh Mussavi Rizi, Conor O’Neill, Jeffrey C Lotz, Paul Anderson, Abel Torres-Espin

**Affiliations:** Department of Computer Science and Software Engineering, California Polytechnic State University, San Luis Obispo, California, USA; School of Public Health Sciences, University of Waterloo, Waterloo, Ontario, Canada; Department of Orthopaedic Surgery, University of California San Francisco, San Francisco, CA, USA; Department of Neurological Surgery, University of California San Francisco, San Francisco, California, USA

**Keywords:** Causal Discovery, Knowledge Graph, Large Language Model, Chronic Lower Back Pain

## Abstract

Causal discovery algorithms are often leveraged for inferring causal relationships and recovering a causal model from data. However, causal discovery from data alone is limited by the structural constraints of the used dataset, the lack of causal logic, and the lack of external knowledge. Thus, data-driven causal discovery can only suggest possible causal relationships at best. To overcome these limitations, Large Language Models (LLMs) and knowledge systems, such as Retrieval-Augmented Generation (RAG), have been proposed as alternatives to data-driven causal discovery and as a method to augment causal discovery algorithms. Using an expert-defined causal graph of chronic lower back pain, we further propose knowledge graph based RAG systems, such as GraphRAG, as an improvement over RAG systems for augmenting causal discovery (F1 0.745), benchmarking its performance against augmenting causal discovery with an LLM (F1 0.636), augmenting causal discovery with RAG (F1 0.714), and causal discovery alone (F1 0.396). We also explore the impact of different prompting methods for causality, such as querying for the plausibility of causal relationships, the presence of statistical associations, and the existence of temporal causal relationships, as inspired by the methodology of the domain experts constructing our ground truth. Lastly, we discuss how applications of LLMs, RAG, and graph-based RAG systems can impact and accelerate the causal modeling of chronic lower back pain by bridging the gap between domain knowledge and data driven approaches to causal modeling.

**Graphical Abstract:** 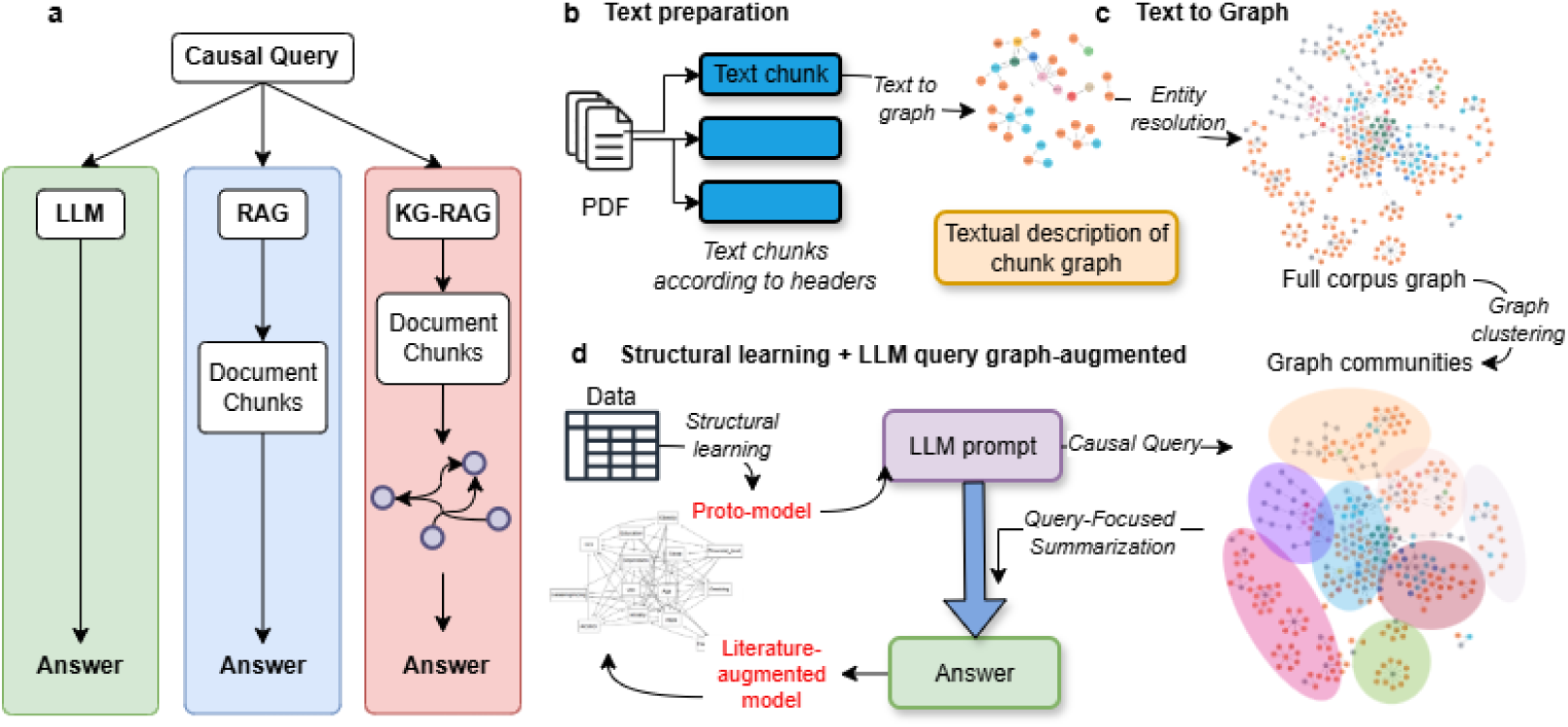

## 1. Introduction

Establishing causal relationships between variables from observational data is critical in health research, particularly in domains where randomized experiments are unfeasible, unethical, or impossible. Causal models provide a formal way to represent the hypothesized causal relations between variables. These models can be used to inform the subsequent analysis, for example, to determine the set of covariates to use for adjustment in a regression analysis to control for confounding factors. Causal models are typically expressed through causal graphs or systems of structural equations. When represented graphically, variables appear as nodes and causal relationships are encoded as directed edges [1].

Causal models can be constructed by experts synthesizing domain knowledge, drawing on evidence from prior studies and expert judgment. However, developing such a model can be an involved process and take considerable time, particularly when many factors must be considered or when the underlying mechanisms are complex and supported by a large and heterogeneous volume of evidence in the literature [2]. Under these conditions, constructing expert causal models that capture the full complexity of a problem becomes impractical, and researchers often reduce the scope to a small handful of variables.

Alternatively, causal models can be constructed using causal discovery methods, a class of data-driven algorithms designed to learn structural models represented as graphs from data [3, 4, 5]. These methods attempt to infer the structure that best captures dependence patterns that may reflect underlying causal relations. Structural learning algorithms are often applied to estimate a graph depicting the true generative process of the data by assessing the presence of directed edges iteratively and relying on statistical assumptions (e.g., conditional independence), heuristics (optimizing a score for fitness to the data), or a combination of both [3, 4, 5].

Causal discovery methods are faster than manual expert causal model construction, scale to large sets of variables, and are derived from observed data directly. However, such methods generally recover only a Markov equivalence class, rather than a uniquely oriented causal graph [6]. More importantly, structural learning methods rely solely on the information present in the data, unless domain knowledge is explicitly encoded, for example, by imposing constraints in the edge search space or by incorporating the prior knowledge by constraining the space of allowable graphs. An alternative to using prior knowledge as search constraints is an expert-in-the-loop approach, in which experts interact with data-driven methods to provide knowledge during the discovery process [7]. However, determining prior constraints or actively interacting with experts in large multi-domain datasets is challenged by the number of domain experts required, limits on the domain expert knowledge, and time efficiency to evaluate all possible evidence in favor or against each potential edge.

To facilitate the study of complex causal structures across diverse areas of health research, we require approaches that retain the scalability of data-driven causal discovery while enabling the rapid and accurate incorporation of domain-expert knowledge. Large language models (LLM)-based causal discovery has been explored as an alternative to traditional expert-driven determination [8, 9, 10, 11]. Here, we propose a novel LLM-based causal discovery system to address these challenges and improve the incorporation of domain knowledge during the causal discovery process. In particular, we focus on chronic low back pain (cLBP) and use a clinically derived, expert-validated causal graph [12] as a gold standard for developing and evaluating our approach.

## 2. Related work

The use of LLMs to assist in the causal discovery process has gained attention in the last few years [9, 10, 11]. Causal relationships between two pairs of variables can be directly queried in LLMs [8], and Kıcıman et al. suggested a search of all possible pairs of variables to complete a full graph [11]. Jiralerspong et al. [13] argued that testing for all possible pairs is unfeasible in a large set of variables. They evaluated a framework using LLMs for full causal graph discovery using only metadata information about the variables of interest and LLM internal knowledge. Their approach first takes a broad search strategy to reduce the search space of pairwise associations, improving the efficiency of causal discovery. In addition, they also investigated the incorporation of observational data into the framework. Cohrs et al., explored the potential of LLMs as surrogates for domain experts in generating causal graphs [14]. They queried LLMs with prompts reflecting a conditional independence question and integrated the response into the PC conditional-independent-based structural learning algorithm [6]. The authors demonstrated that LLMs can effectively complement data-driven methods for causal discovery by testing for conditional independence using an LLM. Ben et al., showcased a two-step process where LLMs are first used to establish a baseline causal structure that is then passed to data-driven algorithms as constraints [15]. Darvariu et al., assessed the efficacy of LLMs in providing prior information for causal graph discovery [16]. Through different prompting strategies, the authors found that LLMs can effectively specify prior causal structures, aiding the causal discovery process. Khatibi et al. [17] described the Autonomous LLM-Augmented Causal Discovery Framework (ALCM). This framework synergizes data-driven causal discovery algorithms with LLMs to automate the generation of accurate and interpretable causal graphs. The study demonstrates that integrating LLMs can address challenges associated with expert knowledge limitations and time efficiency in evaluating constraints.

These previous works use the LLM’s internal knowledge to either query pairwise associations directly, or reason about the conditional independence between a set of variables. However, the efficacy of the methods depends on the LLM used and whether the LLM has sufficient knowledge of the context for which a causal model is to be discovered. Cohrs et al., discussed using Retrieval Augmented Generation (RAG) as a potential approach to improve knowledge relevance [14]. Zhang et al. followed through by combining LLMs and RAG with a large scientific corpus [18]. They propose the LLM assisted causal recovery (LACR) method by first verifying the existence of an edge between each pair of variables using LLMs to query the retrieved scientific documentation, LLM’s internal knowledge, and the output of data-driven methods. This contrasts with directly querying LLMs to reason on the presence of a causal edge. The LACR follows the process by directly querying the LLM for the orientation of the edges discovered during the initial phase. The authors conclude that the process of providing external knowledge to LLM-based causal graph discovery is essential to knowledge updating of the latest research.

Although RAG is instrumental in providing external knowledge that LLMs can use for direct retrieval and reasoning, they experience some limitations, such as effectively answering global questions on an entire corpus [19]. Recent solutions such as GraphRAG propose the structuring of the external documentation as a knowledge graph (KG) that can then be used for supporting LLM reasoning [19]. Following these advances, we propose a novel knowledge-augmented causal discovery system that uses LLMs and KGs to aid data-driven structural learning methods for the task of recovering a causal graph. We evaluate and demonstrate our approach in a real-world case of recovering a causal graph determined by experts in cLBP, and discuss the implications for future methodological research and real-world applications in biomedical research.

## 3. Methods

### 3.1. Study setting and participant data

The study setting and participant data have been previously described [12]. Briefly, we used baseline data from 1,868 participants enrolled in the University of California San Francisco BACKHOME study [20] who completed the baseline survey through April 2023. The BACKHOME is a longitudinal online observational study enrolling patients with cLBP across the United States [20]. The online baseline survey included questions on demographics, back pain, pain impact on quality of life, pain beliefs, back pain treatment, medications, medical history, health habits, and traumatic experiences.

### 3.2. Expert-derived causal graph

A causal graph for the study cohort was previously developed [12] through expertise using a rigorous evidence synthesis for constructing directed acyclic graphs (ESC-DAG) approach [21]. The study aimed to determine causal risk factors for five exposures previously identified in Mendelian randomization research on pain outcomes. The pain outcome was the Pain, Enjoyment of life, and General activity (PEG) scale, a three-item scale for assessing pain intensity and interference [22]. The five exposures were alcohol use, smoking, sleep disturbance, depression, and obesity. A causal graph was constructed using ESC-DAG and evidence from the literature to clarify the causal structure, identify confounders of the relationship between each exposure of interest and PEG, and define adjustment sets of covariates for regression analysis of the exposure and outcome associations (Figure 1). The construction and analysis of the causal graph is described in detail elsewhere [12]. In this work, we use the causal graph determined by the experts as ground truth to be recovered through our knowledge-augmented causal discovery framework. Note that the expert-derived causal graph we are using is not a DAG, and it contains cycles. We have chosen this as a use case because cLBP is a leading cause of disability in the world [1] with significant clinical and public health interest, the graph is of a manageable size (14 variables), the set of articles to use as background knowledge is well-defined, we have expertise in the domain, we are part of the BACKHOME team, and have access to the underlying observed data.

**Figure 1:**
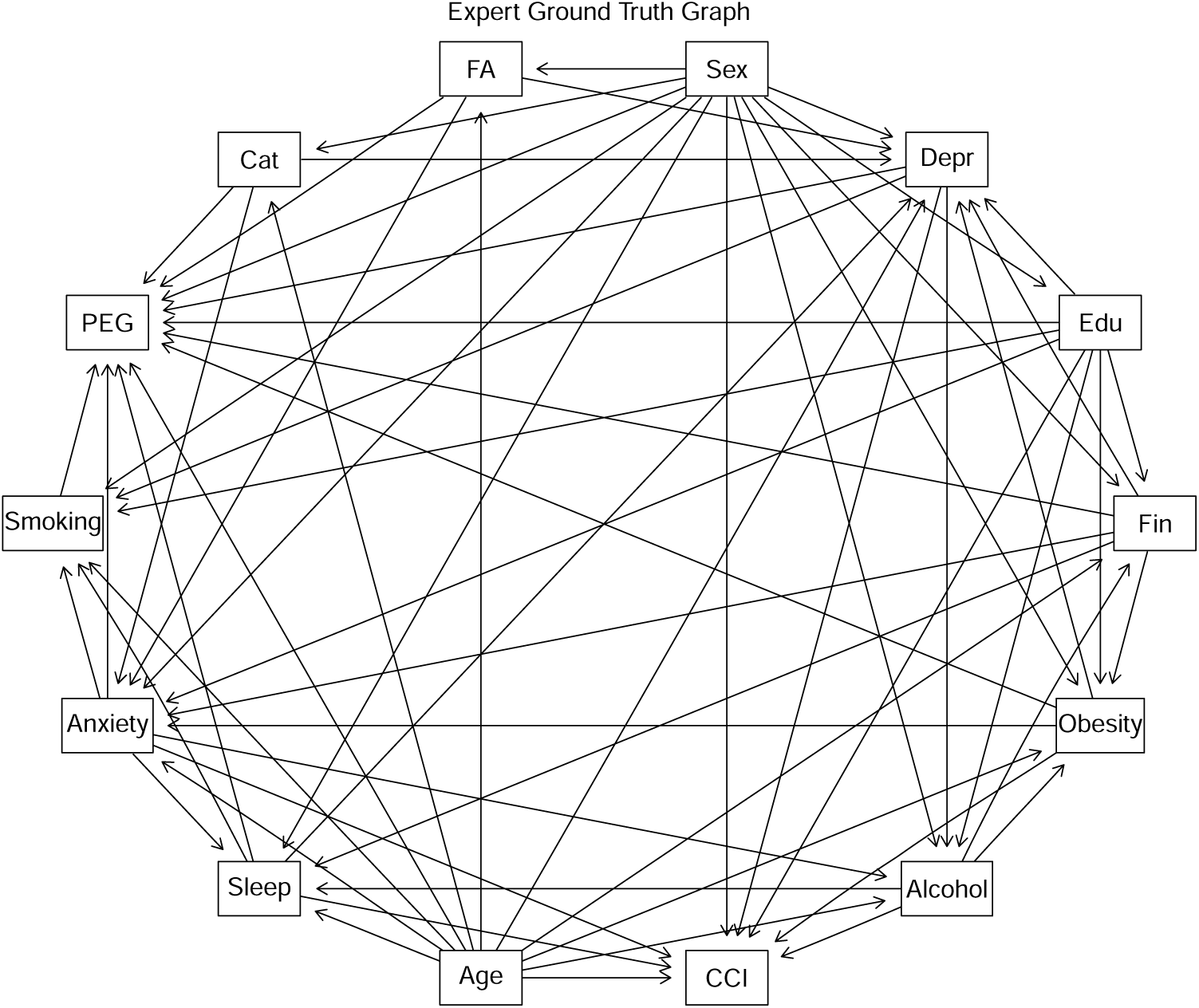
Expert ground truth causal graph showing causal relationships between variables. Abbreviated nodes include: FA for Fear Avoidance, Cat for Pain Catastrophizing, Sleep for Sleep disturbance, Fin for Financial Level, Edu for Education, Depr for Depression, and CCI for Charlson Comorbidity Index. Other nodes display full variable names.

### 3.3. Knowledge systems

Three knowledge systems were developed for this work with increased complexity: A system with a vanilla LLM, a system with the same LLM + a RAG, and our proposed LLM + KG-RAG (leveraging Knowledge Graphs to retrieve context for LLM systems) (Fig. 2a).

**Figure 2:**
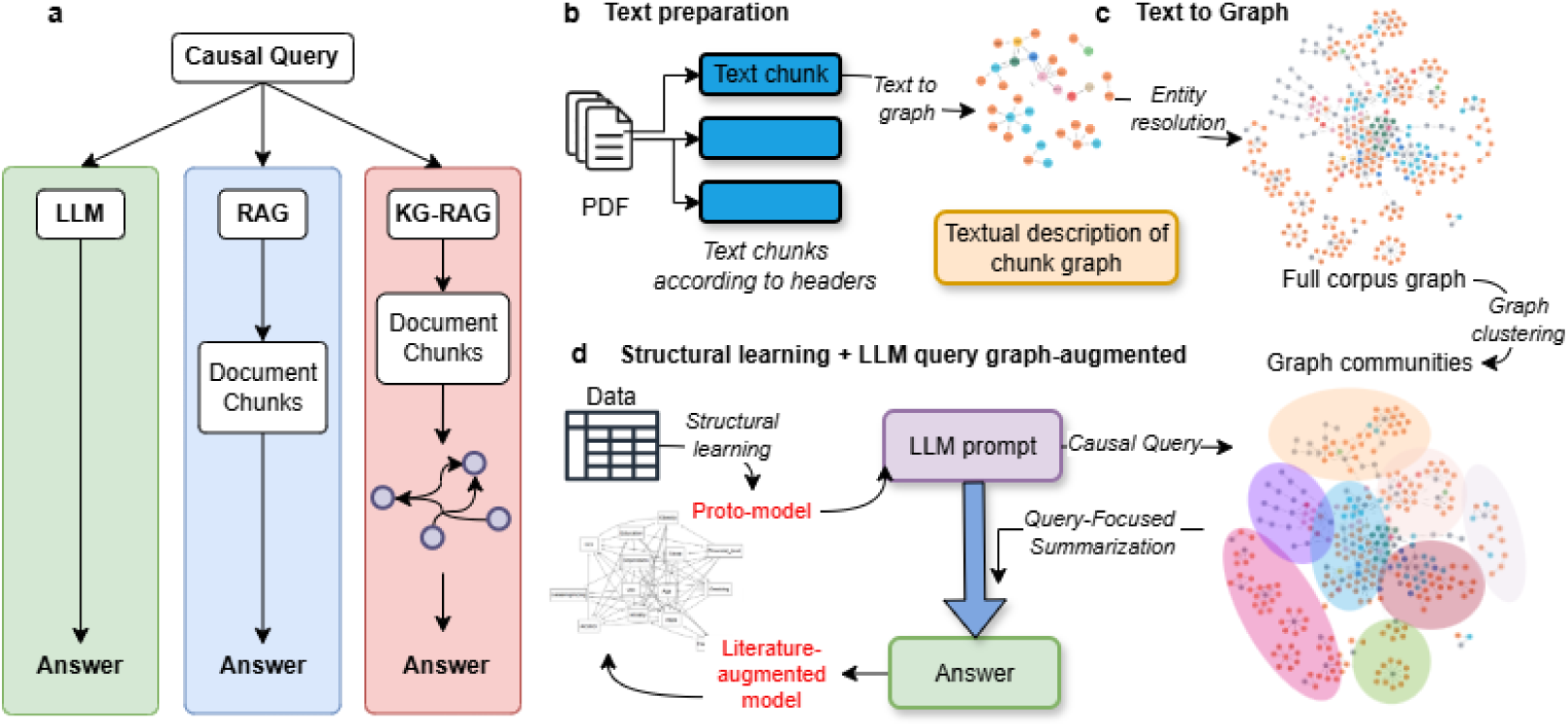
Knowledge systems. (a) Shows the three knowledge systems that we compared. (b) to (d) shows a diagram of the main steps for the KG-RAG system construction. The system starts by parsing and chunking the documents (b), then building a KG (c) followed by graph clustering to find concept communities. (d) illustrates the concept of augmenting structural learning with causal queries to the KG.

#### 3.3.1. KG-RAG knowledge system

To serve as an external knowledge source for our LLM, we constructed a knowledge graph (KG) from the same source documents used by domain experts, drawing on the KG construction methods introduced in Edge et al. [19], as shown in Figure 2. First, we manually collected the papers used by the experts to build the causal graph as PDFs and converted them to markdown files using Marker^2^. Afterwards, we clean the text by filtering sections of the paper outside of the main body, such as acknowledgments, supplementary info, funding disclosures, references, etc, as well as citations. Then, we split the markdown files into 600 token text chunks, with a 100 token overlap, and prompt the LLM to extract all of the entities and relationships present, as done in GraphRAG. We use a prompt originally in the GraphRAG system that we tuned to our use case, and the Python library Outlines [23] to constrain the generation of the LLM, so that only Entities and Relationships are generated. Notably, we do not provide any examples to the LLM (zero-shot prompting), and we conduct only a single pass over the text chunks. Post entity and relationship extraction, we store the KG elements into Neo4j, a graph database. After, we perform entity resolution, following a three-step process [24]. First, for each entity, we perform a nearest neighbor search based on semantic similarity, using embeddings generated by the embedding model, S-PubMedBert-MS-MARCO [25]. Second, from the nearest neighbors, we filter out syntactically dissimilar entities, using edit distance. Third, we prompt the LLM to pass over each group of nearest neighbors and distinguish groups of entities identical in meaning. After identifying groups of entities to be merged, we join the nodes within Neo4j. Finally, we cluster the entities and relationships extracted by the LLM into hierarchical communities using the Leiden Algorithm [26]. For each community, we used the LLM to summarize all of the entities and relationships within the cluster following the methods described in GraphRAG, forming community-wide natural language summaries for downstream analysis. Similarly to entity and relationship extraction, we prompt the LLM for community summarization by tuning a prompt used in the original GraphRAG implementation.

After the KG has been constructed, we augment queries to the LLM with relevant information retrieved from the KG, following the local search method also introduced in Microsoft’s GraphRAG [19], as shown in Figure 3. We limit the quantity of relevant context retrieved from the KG to 8000 tokens, which was empirically determined in the GraphRAG paper to be optimal. In addition, we also provide the definitions for the variables in our dataset, as used by the domain experts (see Appendix C for more details). Prior to the causal queries, we inject the context into the prompt.

**Figure 3:**
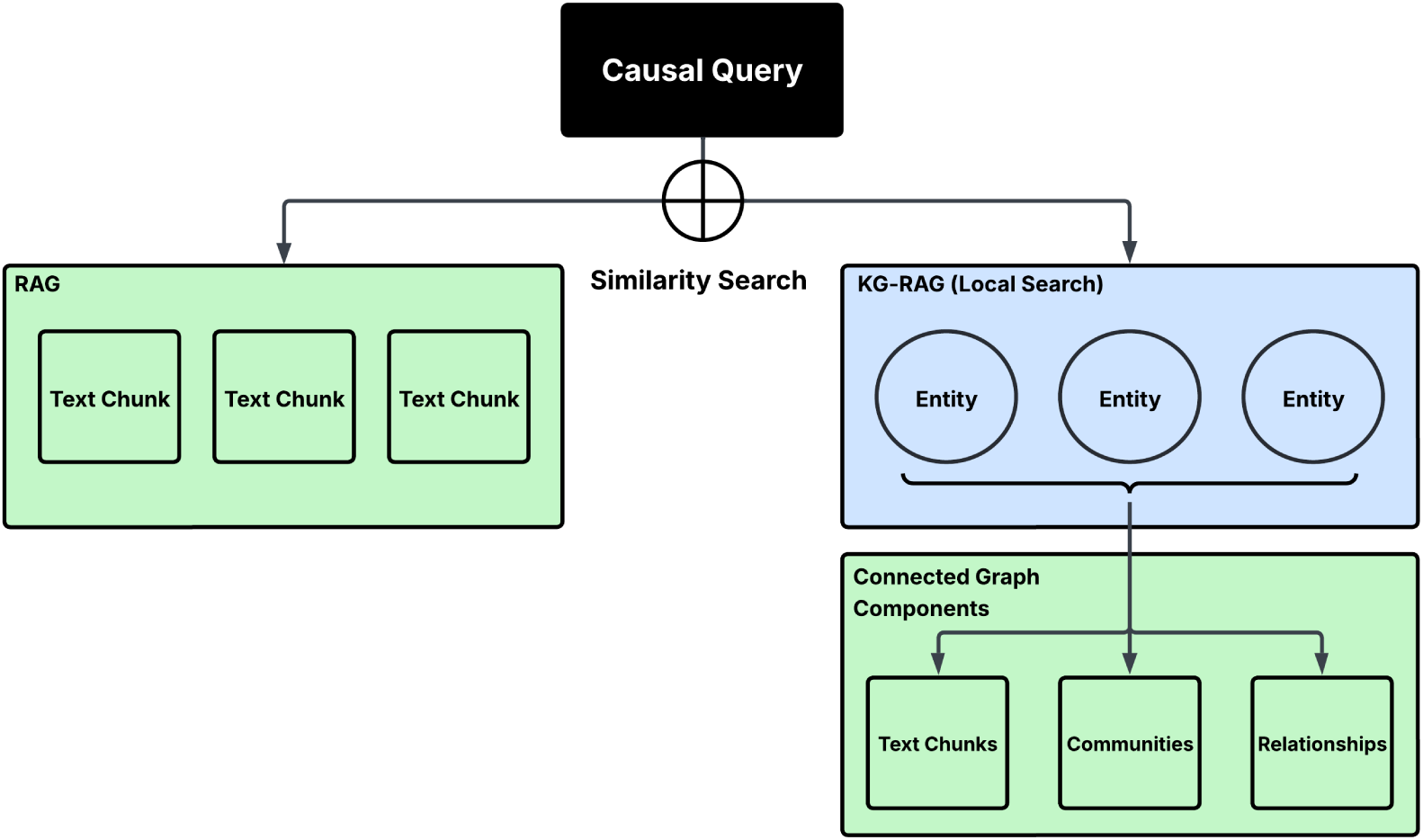
Comparison between KG-RAG (Local Search) and RAG methods. RAG leverages vector similarity search to identify text chunks similar to the query. KG-RAG leverages vector similarity search to identify entities similar to the query, retrieving the entities and components such as the summaries of communities the entities belong to, the relationships attached to the entities, and text chunks where the entities were extracted from.

#### 3.3.2. Comparisons with LLM and RAG-based knowledge systems

To benchmark the effectiveness and utility of the KG-RAG method, we evaluate the two other knowledge systems: using the LLM without a knowledge retrieval mechanism, and using the LLM within a standard RAG system where relevant text chunks from the documents used in the expert-driven literature review are retrieved via vector similarity search over embeddings, as shown in Figure 3. For the standard RAG comparison, we convert the papers into text chunks as described in the knowledge system. Then, we embed each of the text chunks using the embedding model S-PubMedBert-MS-MARCO, and, afterwards, we store all chunks in a vector index using FAISS [27]. When retrieving relevant text chunks during query time, we follow a similar method as described in the knowledge system, where we limit the amount of relevant information supplied to the LLM to a context window of 8000 tokens. Similarly to the KG-RAG system, we provide the definitions of each variable, as used by the domain experts when determining the ground truth, when querying the LLM with retrieved knowledge and the LLM without retrieved knowledge.

#### 3.3.3. System-Wide parameters

Across KG-RAG, LLM, and RAG systems, we use Mistral 7B-Instruct v0.3 [28]. We selected Mistral 7B-Instruct v0.3 as our LLM because it is open source, providing fine-grained control over system prompts and hyperparameters, while controlling for unknown variables characteristic of proprietary systems. Furthermore, since Mistral 7B-Instruct v0.3^3^ was released prior to the publication of the expert causal graph [12], the ground truth could not have been included in the training data of the LLM.

In addition, to improve the performance of the LLM and add additional insight into how the LLM is reasoning through the causal queries, we use chain of thought querying [29] for each of our knowledge systems.

### 3.4. Causal prompts

During the literature review completed in the process of constructing the expert-driven causal graph, the domain experts noted whether each of the pairwise associations was plausible, had a known association, and had a known temporal relationship [12]. Inspired by their methodology and to investigate different prompt strategies for querying LLMs about causal relationships, for each pairwise association posed to the knowledge systems, we make three independent zero-shot direct queries:

#### 3.4.1. Plausibility query

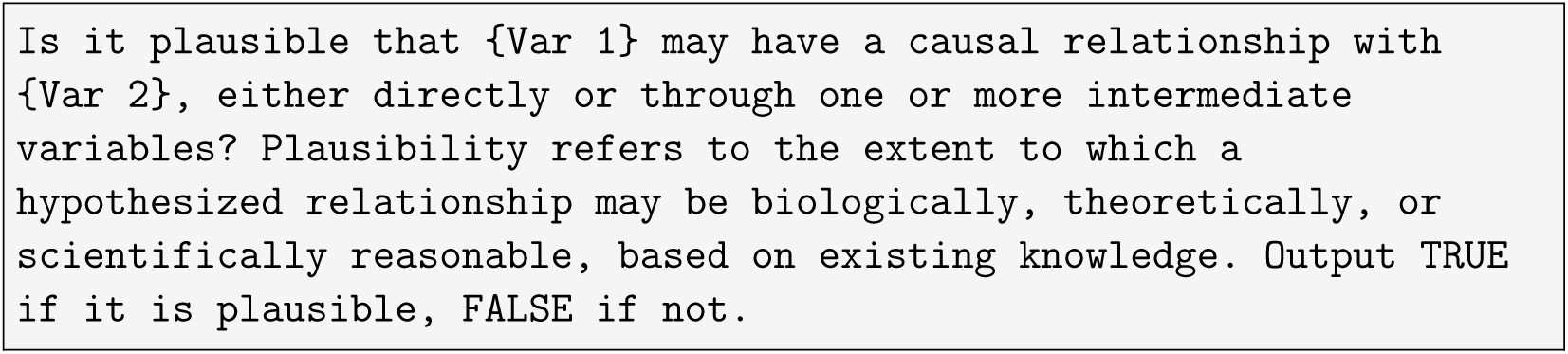

#### 3.4.2. Association query

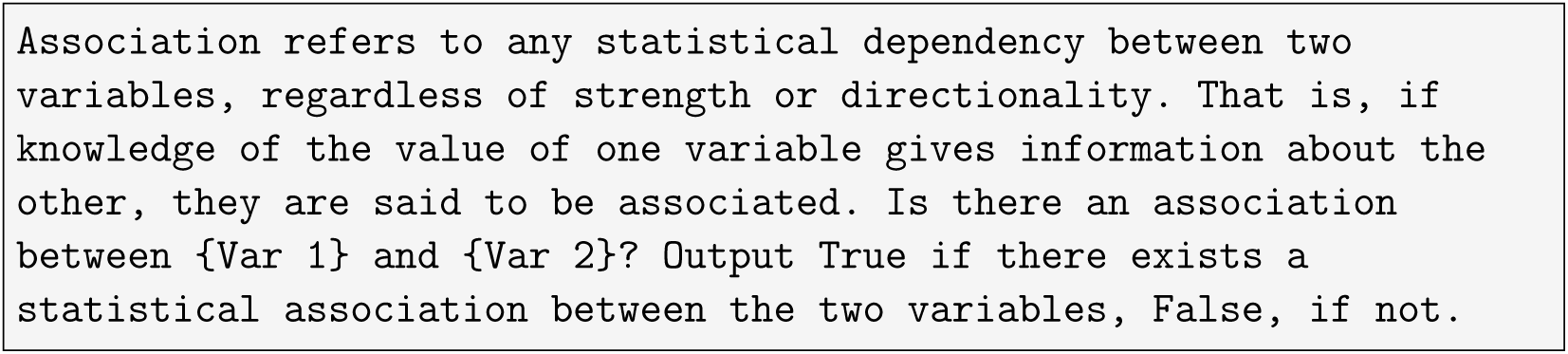

#### 3.4.3. Temporality query

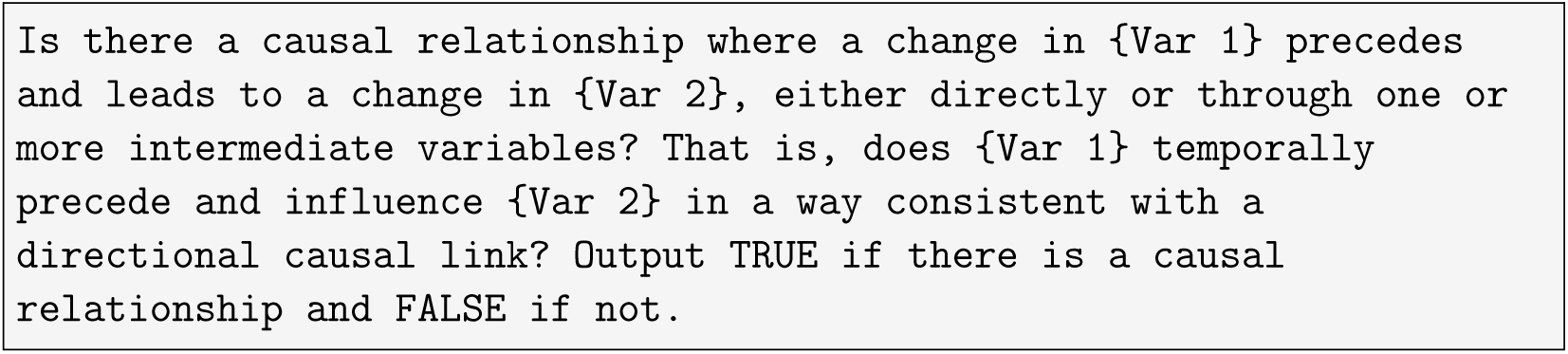

#### 3.4.4. Direct causal query

In addition, we investigated a form of a direct causal query used by Jiralerspong [13], which is one of the most used forms of queries in the literature. We use the direct causal query for completeness and reference to other previously investigated causal queries.

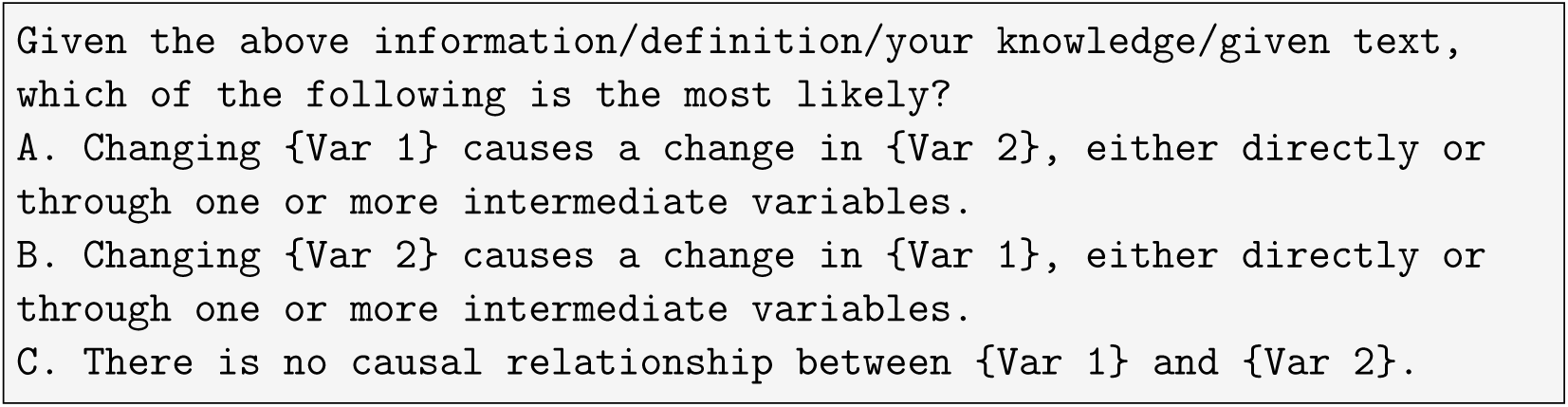

### 3.5. Two-phased causal querying

Similarly to methods such as LACR [18], we split our causal querying process into two phases: (1) determining edge presence and (2) determining edge direction, as shown in Figure 4.

**Figure 4:**
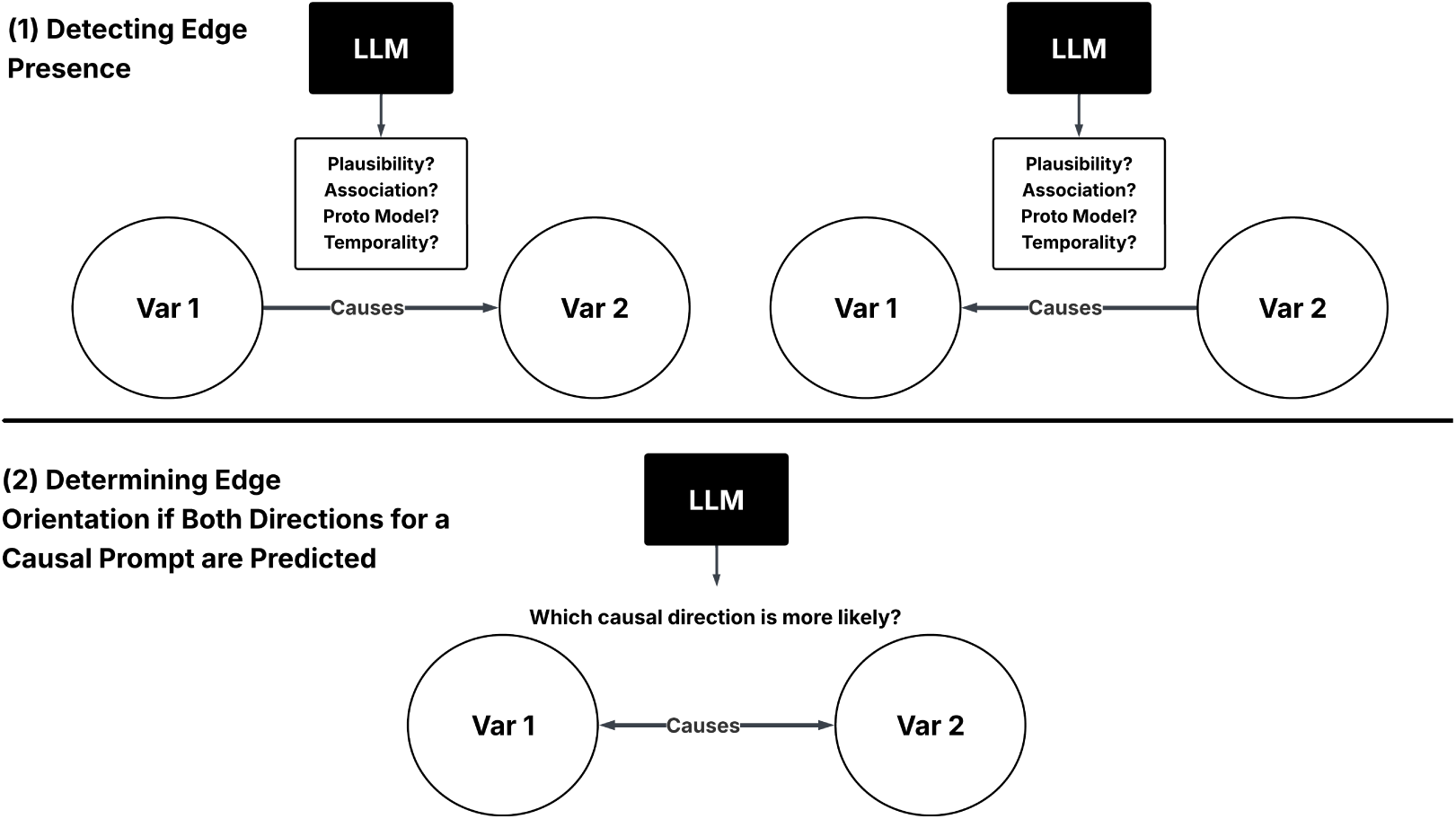
Two Stages of Causal Querying.

#### 3.5.1. Determining edge presence

To determine the presence of an edge between two variables, A and B, we apply Plausibility, Association, and Temporality queries to every possible edge and direction (querying A *→* B and B *→* A), with the exception that no variable can cause Sex and Age, and PEG cannot be a cause with respect to any variable, since PEG was the original outcome of the ground truth. Across all queries, if the knowledge system predicts either direction of an edge to be true, we predict that an edge exists.

#### 3.5.2. Determining edge direction

After determining that an edge exists, we determine the orientation of the edge. First, for each causal prompt, if the knowledge system predicts that a relationship exists for a single direction, then that direction is final. However, if the knowledge system predicts that a relationship exists for both orientations (a bidirectionally predicted edge), we make a second query to determine which direction of causality is more likely to be true, drawing on the prompting strategies introduced in Kıcıman et al [11]. The result of the second query becomes the final prediction for that causal prompt.

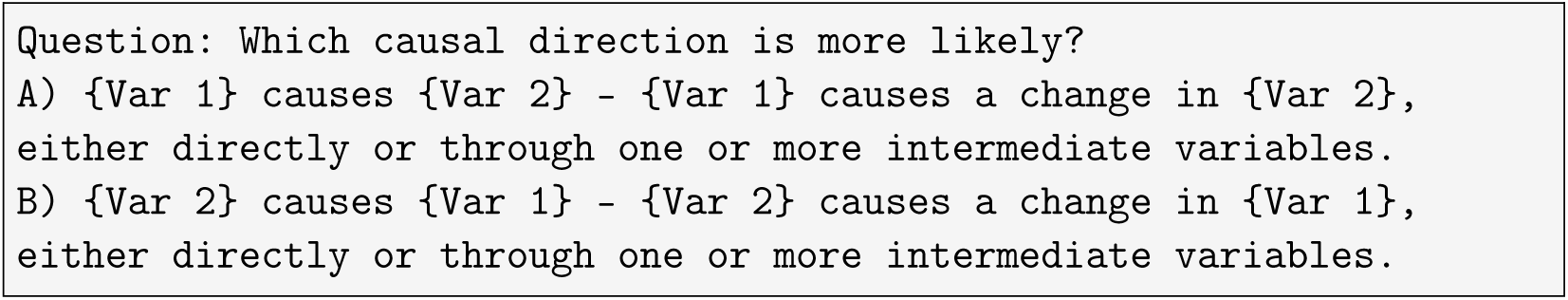

Note, we do not resolve bidirectional edges for the Direct Causal Query, since the Direct Causal Query determines edge presence and edge orientation in the same prompt.

### 3.6. Structural learning

Our setup is agnostic to the structural learning algorithm used for data-driven causal discovery, providing flexibility in choosing methods to determine the proto-model. In this work, we use the hill climb algorithm, a score-based approach to learn the structure from data, as implemented in the bnlearn R package [30], and a custom scoring function. Given that the dataset is a mix of variable types, we used a scoring function in which the local distribution of each node is modeled using a generalized linear model (GLM) and Bayesian Information Criterion (BIC) is used as a score metric (as suggested in the bnlearn documentation). If the child node introduced by a directed edge in the hill climb search is continuous, a linear regression model was used; if it is binary, a logistic regression was used; and if it is categorical, a multinomial regression was used. We bootstrapped the hill climb algorithm 500 times to calculate an edge’s empirical presence strength and direction strength implemented in bnlearn following [31]. Only the edges with strength above a significant threshold as determined by [32] were retained on an averaged consensus graph. This constitutes the data-driven proto-model. The moral graphs of the hill climb results and proto-model were also extracted. The moralized graph is the equivalent of an undirected graph to a directed graph by “marrying the parents” with common children with undirected edges, plus dropping directionality in all edges.

### 3.7. Knowledge-augmented causal discovery

Our framework is similar to previous approaches [17] in which a proto-causal graph is first learned from observed data using structural learning algorithms, and it is then augmented and refined with knowledge extracted from a set of predefined research articles using our knowledge systems (Fig. 2d). We refer to the first data-driven graph as a proto-model, as in general, structural learning algorithms recover a graph capturing the data-generating process up to a Markov equivalent set under the assumption of causal sufficiency and faithfulness [8].

Independent of the structural learning algorithms, we query the LLMs regarding the presence of each possible edge. For Knowledge-Augmented Causal Discovery, we integrate the predictions of the proto-model into our causal querying framework by leveraging the structural learning algorithm as an additional indicator for detecting edge presence, as shown in Figure 5. After taking the union of the edges predicted by the knowledge system and the edges predicted by the proto model, we resolve bidirectionally predicted edges, edges where both directions have been predicted, by querying the LLM to determine which direction of causality is more likely, as done in the second step of the causal querying framework (Sec. 3.5.2).

**Figure 5:**
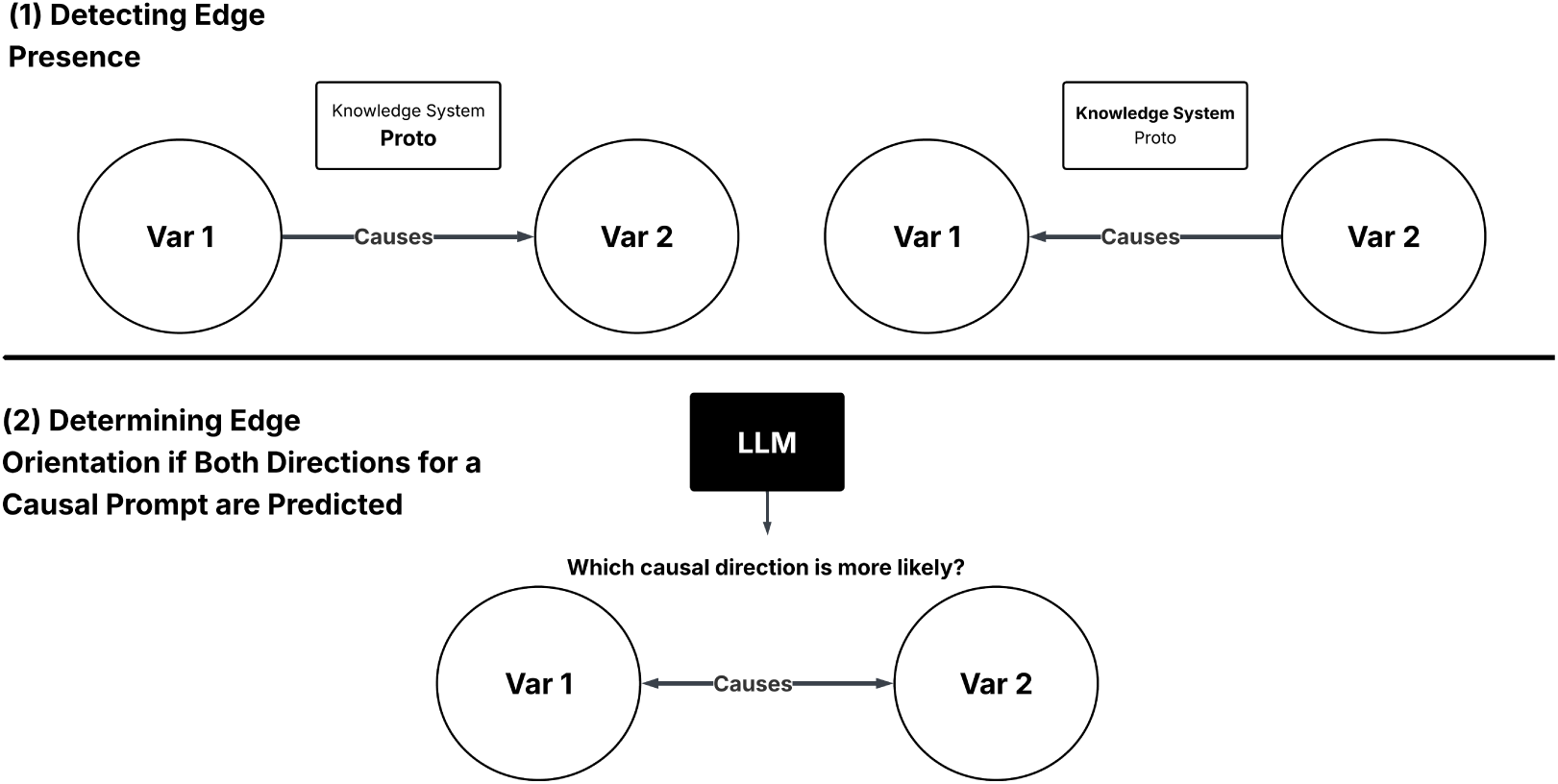
To refine the proto model with the knowledge systems, the edges predicted by the proto model and the edges predicted by the knowledge systems are first combined into a single graph. If both directions of any edge are predicted after taking the union of the knowledge systems and the proto model, a second query is made to determine the correct direction.

### 3.8. Evaluation

We evaluated different knowledge-augmented causal discovery framework settings on their capacity to recover the expert-derived causal graph. For ev-ery graph tested against the expert-derived causal graph, the true positive, false positive, and false negative edges were calculated. In our evaluation, a true positive is a predicted directed edge that is present in the ground truth graph, a false positive is a predicted directed edge that is not present in the ground truth graph, and a false negative is a directed edge in the ground truth graph that is not predicted. As performance metrics, we compute the false discovery rate (FDR), the true positive rate (TPR), and the F1-Score as measures of edge recovery accuracy with respect to the expert-derived graph. In addition, we calculated the Hamming Distance (HD) and Structural Hamming Distance (SHD) between the graph provided by each method and the expert-derived causal graph. The HD measures the distance between the skeletons of two graphs by counting the number of edges that are present in one graph but absent in the other. SHD is a measure of global conformance between two graphs measured by counting the number of edge insertions, deletions, or flips required to transform one graph into another by comparing the adjacency matrices [6]. Lower values of HD and SHD imply higher structural similarity (less differences) between graphs. For completeness, we evaluate randomly generated graphs (100 random graphs) from the structure of the expert-derived causal graph but with random edges, which provides random guessing performance. We also evaluate our knowledge systems alone (LLM, RAG, and KG-RAG without the proto-model), the proto-model alone, and the combination of proto-model augmented using our knowledge system.

## 4. Results

### 4.1. Proto model

For the proto model, the optimal threshold for determining the presence of an edge based on the strength indicated by the hill climb algorithm was 0.538. As the threshold increases, the performance of the consensus model, determined by the 500 bootstrapped iterations of the hill climb algorithm, decreases in F1 score when compared to the expert-driven graph (Fig. 7).

We also compared the moralized consensus graph (the equivalent undirected graph from the hill climb consensus) from the structural learning algorithm for each edge strength threshold with the moralized expert-derived graph. This provides a comparison of the underlying pairwise association structure without considering the edge directions. As shown in Figure 7, the hill climb algorithm produces a much closer structure to the structure of the expert-derived graph when directionality is not considered. This is somewhat expected, as determining pairwise associations is a much easier task than deciding on edge direction, especially in a cross-sectional setting. This suggests that the causal relationships determined by the domain experts are aligned with what can be observed in the data. The resulting proto-model is shown in Fig. 6.

**Figure 6:**
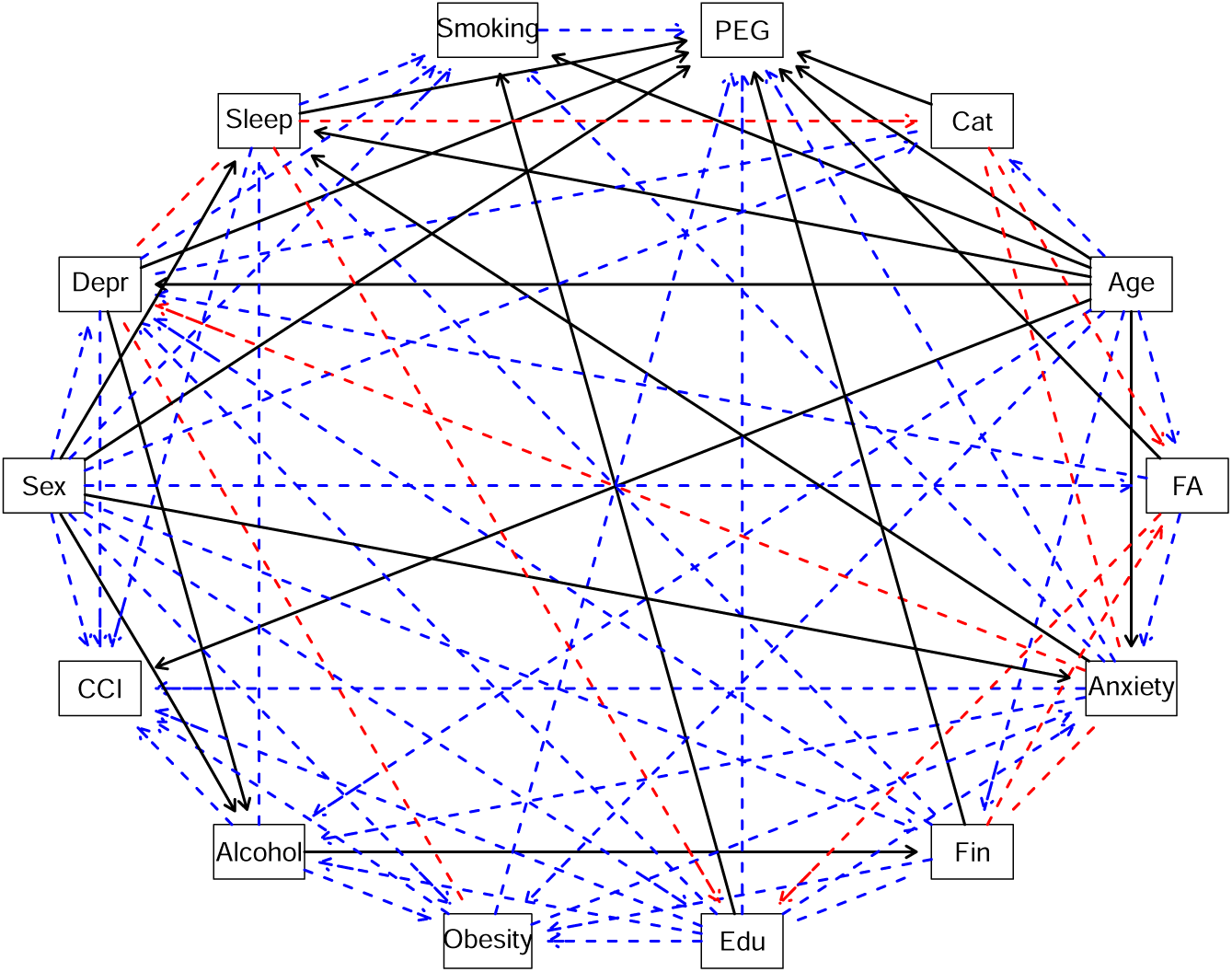
The proto causal graph model. Abbreviated nodes include: FA for Fear Avoidance, Cat for Pain Catastrophizing, Sleep for Sleep disturbance, Fin for Financial Level, Edu for Education, Depr for Depression, and CCI for the Charlson Comorbidity Index. Other nodes display full variable names. Solid black lines denote true positives, dashed red lines denote false positives, and dashed blue lines represent false negatives.

**Figure 7:**
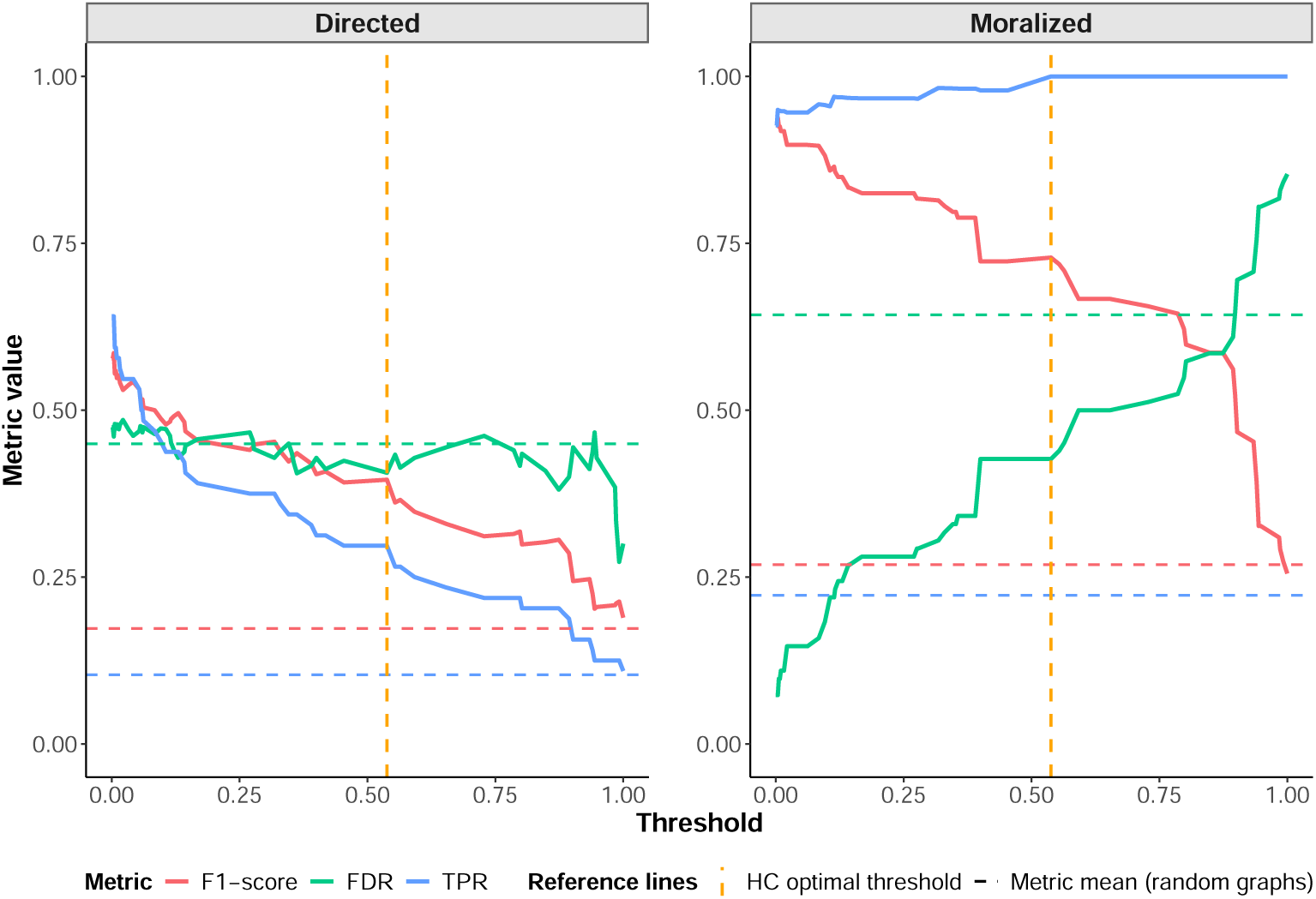
Performance of the structural learning. The performance metrics (F1-score, FDR, and TPR by increasing the strength threshold are shown for the consensus hill climb graph over 500 bootstraps (left), and the corresponding moralized graph (right). Dashed horizontal lines represent the average of each metric over 100 random graphs (same color as the corresponding metric), while the dashed vertical line indicates the optimal threshold of the HC algorithm (0.538).

We then compared the resulting proto-model graph model against a benchmark of randomly generated graphs (Table. 1).

**Table 1:**
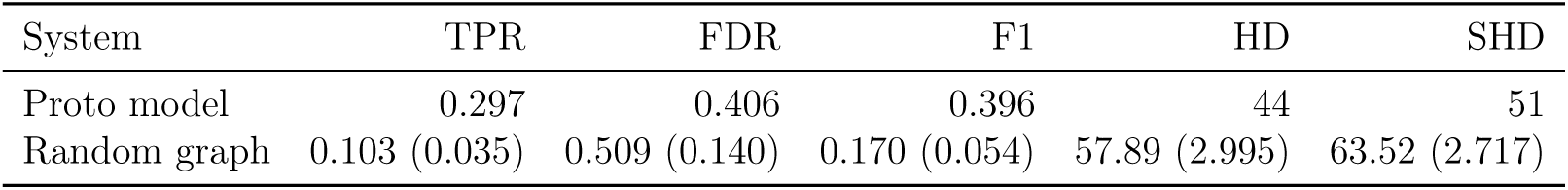
Comparison of the proto model’s performance at the optimal threshold selected by the HC algorithm, evaluated against 100 randomly generated graphs. Standard deviations of each metric for random graphs are reported in parentheses.

As shown in Table. 1, the proto model outperforms the randomly generated graphs with an F1-Score of 0.396 as opposed to 0.170. Notably, the TPR of the proto model is much higher than the TPR of the randomly generated graphs (0.297 vs. 0.103), and the FDR of the proto model is much lower (0.406 vs. 0.509). The SHD for the proto model is also lower in comparison to the randomly generated graphs (51 vs. 63.5).

### 4.2. Knowledge systems

In this section, we describe the performance of the LLM, RAG and KG-RAG knowledge systems. Table 2 shows the performance of each knowledge system across every metric prior to resolving bidirectional edges. Each knowledge system achieves their best performance when predicting Plausibility, with LLM reaching an F1 of 0.627, RAG scoring an F1 of 0.612, and KG-RAG achieving an F1 of 0.602. Notably, the performance across all knowledge systems decreases across Plausibility, Association, and Temporality. In comparison to the Direct Causal Prompt, all systems achieve a higher overall performance when predicting Plausibility, Association, and Temporality, with the exception of the LLM with Temporality and KG-RAG with Temporality.

**Table 2:** Performance of each knowledge system prior to resolving bidirectional edges.

| **System** | **Metric** | **TPR** | **FDR** | **F1** | **HD** | **SHD** |
| --- | --- | --- | --- | --- | --- | --- |
| LLM | Plausibility | 1.000 | 0.543 | 0.627 | 26 | 57 |
| LLM | Association | 0.516 | 0.554 | 0.478 | 40 | 64 |
| LLM | Temporality | 0.109 | 0.563 | 0.175 | 56 | 61 |
| LLM | Direct Causal | 0.188 | 0.571 | 0.261 | 52 | 61 |
| RAG | Plausibility | 0.984 | 0.556 | 0.612 | 27 | 59 |
| RAG | Association | 0.641 | 0.506 | 0.558 | 34 | 59 |
| RAG | Temporality | 0.359 | 0.489 | 0.422 | 42 | 55 |
| RAG | Direct Causal | 0.313 | 0.487 | 0.388 | 46 | 55 |
| KG-RAG | Plausibility | 0.969 | 0.563 | 0.602 | 28 | 60 |
| KG-RAG | Association | 0.391 | 0.583 | 0.403 | 46 | 66 |
| KG-RAG | Temporality | 0.172 | 0.500 | 0.256 | 50 | 58 |
| KG-RAG | Direct Causal | 0.234 | 0.483 | 0.323 | 48 | 58 |

Table 3 shows the performance of each knowledge system and metric after resolving edges predicted to be bidirectional. Each knowledge system achieves its highest performance predicting Plausibility, with KG-RAG (0.737) outperforming the RAG (0.706) and LLM (0.636) systems. Notably, the SHD of each system and metric decreases after resolving bidirectionality when compared to Table 2.

**Table 3:**
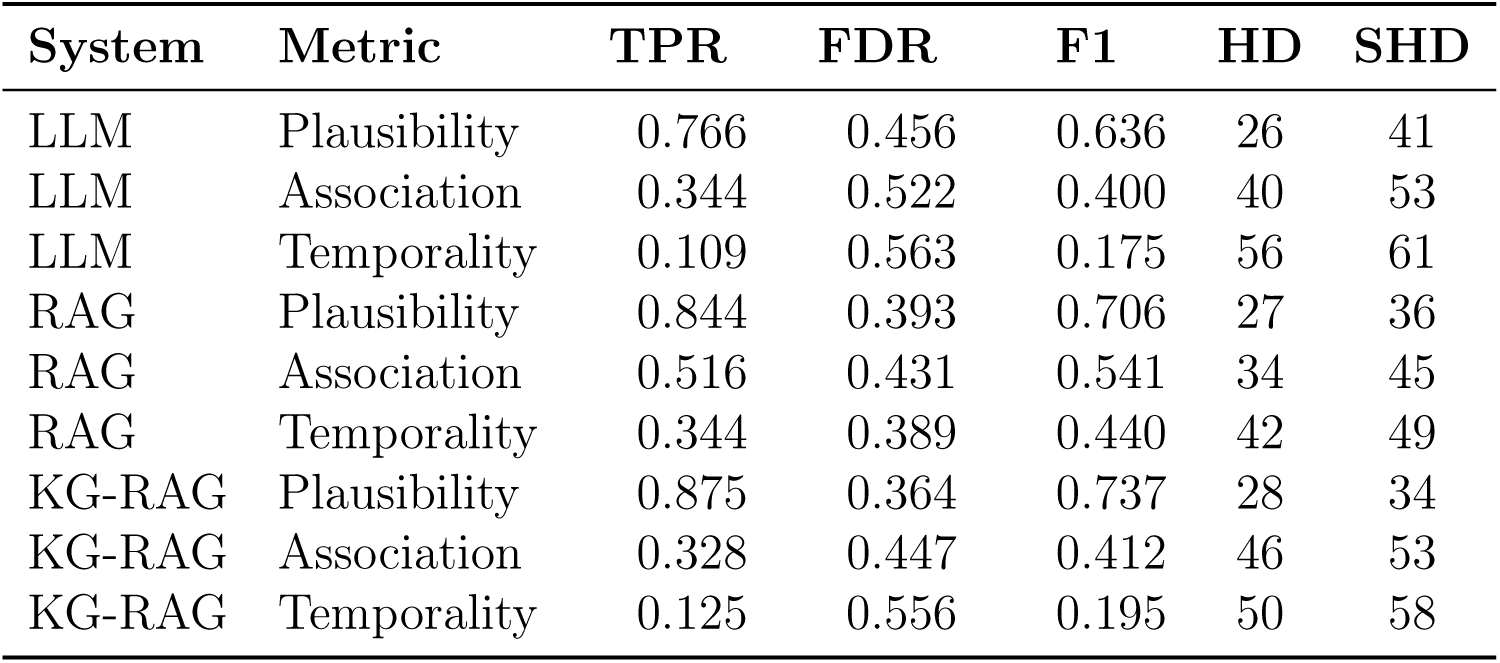
Performance of each knowledge system after resolving bidirectional edges.

### 4.3. Knowledge-augmented causal discovery

Before augmenting the structural learning algorithm with the predictions of the knowledge systems, we first analyze the alignment between the knowledge systems and the causal discovery algorithm (Table 4). To measure the agreement between knowledge systems and the proto model, we measure the HD and SHD between the predictions of the knowledge systems and the proto model. Notably, across all knowledge systems, the structural learning algorithm is least aligned with Plausibility, with the highest HD and SHD in comparison to Association, Temporality, and the Direct Causal Prompt.

**Table 4:** The HD and SHD between each knowledge system and metric with respect to the proto-model. For Plausibility, Association, and Temporality, the causal graphs after resolving bidirectionally predicted edges (Table 3) were compared with the proto model.

| System | Metric | HD | SHD |
| --- | --- | --- | --- |
| LLM | Plausibility | 58 | 66 |
| LLM | Association | 46 | 52 |
| LLM | Temporality | 36 | 37 |
| LLM | Direct Causal | 36 | 43 |
| RAG | Plausibility | 59 | 69 |
| RAG | Association | 50 | 58 |
| RAG | Temporality | 38 | 46 |
| RAG | Direct Causal | 46 | 51 |
| KG-RAG | Plausibility | 58 | 69 |
| KG-RAG | Association | 42 | 48 |
| KG-RAG | Temporality | 34 | 36 |
| KG-RAG | Direct Causal | 32 | 38 |

Following our results above suggesting that the structural learning algorithm fails mostly in edge orientation but succeeds in capturing the underlying association structure, we explore augmenting the output of the hill climb algorithm (the proto model) with the predictions of the knowledge systems, as described in Figure 4.

Table 5 displays the performance of augmenting the proto model with every knowledge system and metric after resolving bidirectionally predicted edges. Overall, as shown in the Δ F1*_p_*, adding any knowledge system and metric increases the performance relative to the proto model alone. Furthermore, the knowledge systems least aligned with the proto model (higher HD and SHD values), such as LLM with Plausibility, RAG with Plausibility, and KG-RAG with Plausibility (Table 4), lead to the largest increases in overall performance relative to the proto model. Moreover, as shown in the Δ F1*_ks_*, combining the proto model with the knowledge system improves overall performance, with the exception of LLM with Plausibility. Notably, both Δ F1*_p_* and Δ F1*_ks_* are all zero or positive, indicating that combining both the proto model and the knowledge systems produces a prediction that is at least as performant or better than either method individually.

**Table 5:** The proto model combined with each knowledge system after resolving bidirectionality. Δ F1*_p_* refers to the difference in F1 score when compared to the proto-model. Δ F1*_ks_* refers to the difference in F1 score when compared to the knowledge systems. For Plausibility, Association, and Temporality, we compare with the results after resolving bidirectionally predicted edges (Table 3).

| System | Metric | TPR | FDR | F1 | $\Delta F1_p$ | $\Delta F1_{ks}$ | HD | SHD |
| --- | --- | --- | --- | --- | --- | --- | --- | --- |
| LLM | Plausibility | 0.766 | 0.456 | 0.636 | 0.241 | 0.000 | 26 | 41 |
| LLM | Association | 0.547 | 0.435 | 0.556 | 0.160 | 0.156 | 30 | 43 |
| LLM | Temporality | 0.359 | 0.452 | 0.434 | 0.038 | 0.259 | 38 | 49 |
| LLM | Direct Causal | 0.406 | 0.447 | 0.468 | 0.073 | 0.208 | 37 | 48 |
| RAG | Plausibility | 0.859 | 0.389 | 0.714 | 0.318 | 0.008 | 26 | 35 |
| RAG | Association | 0.656 | 0.400 | 0.627 | 0.231 | 0.086 | 28 | 39 |
| RAG | Temporality | 0.516 | 0.377 | 0.564 | 0.168 | 0.124 | 35 | 43 |
| RAG | Direct Causal | 0.516 | 0.431 | 0.541 | 0.145 | 0.153 | 34 | 45 |
| KG-RAG | Plausibility | 0.891 | 0.360 | 0.745 | 0.349 | 0.008 | 27 | 33 |
| KG-RAG | Association | 0.531 | 0.393 | 0.567 | 0.171 | 0.155 | 32 | 42 |
| KG-RAG | Temporality | 0.359 | 0.452 | 0.434 | 0.038 | 0.239 | 36 | 48 |
| KG-RAG | Direct Causal | 0.422 | 0.372 | 0.505 | 0.109 | 0.182 | 35 | 44 |

## 5. Discussion

In this work, we developed and evaluated knowledge systems to perform knowledge-augmented causal discovery. These methods can aid in building causal graphs by combining data-driven approaches with LLMs leveraging their internal knowledge, external documents, and/or insights extracted from a KG to reason over causal relationships. Our results suggest that KG-RAG can effectively propose plausible causal relationships, and combining these knowledge systems with causal discovery algorithms can provide insight into a larger range of variables and relationships. Furthermore, the results support the application of LLM-driven knowledge systems to aid in the causal modeling of cLBP through unifying domain knowledge and data driven analysis.

### 5.1. Prompt methodology and causal querying

Motivated by how the experts built the ground truth causal model, we queried the LLMs to determine three different metrics: whether the relationship is plausible, whether there exists a statistical association, and whether the causal relationship possesses a known temporal aspect (evidence that the cause occurred before the effect). Unlike prior approaches documented in the literature, our prompts focus on determining particular qualities of a causal relationship (i.e Temporality, Association, Plausibility), as opposed to leveraging LLMs for systematically performing statistical association tests [18] [14] or for making direct determinations of causality [11]. For completeness, we also show our results using a direct causal prompt well-known in the literature [13].

Given the decreasing performance across Plausibility, Association, and Temporality across knowledge systems, as shown in Tables 2 and 3, the results may indicate that LLMs can effectively suggest possible causal relationships, but can struggle to decisively and correctly determine associations and causality. This suggests that the future application of LLMs and knowledge systems to causal modeling should be restricted to support roles, aiding algorithms and humans alike by proposing possible relationships as opposed to completely determining causality independently.

Furthermore, with the exception of RAG with Temporality and KG-RAG with Temporality, all knowledge systems and metrics (both before and after resolving bidirectionally predicted edges) outperform the knowledge systems when prompted with the Direct Causal prompt, as shown in Table 2 and Table 3. This may indicate that determining edge existence and orientation should be separated into two distinct steps, as similarly done in LACR [18].

### 5.2. Comparisons between knowledge systems

As shown in Table 3, utilizing RAG and KG-RAG can improve the overall performance of LLMs for causal modeling. Zhang et al [18] similarly applied RAG to LLM-driven causal modeling in the LACR framework. However, in the LACR study, RAG did not introduce immediate improvements to their results, since the authors found discrepancies between the ground truth and the retrieved literature, leading the LLM to make incorrect predictions until the authors updated their ground truth to reflect the latest research. Since the documents used in the RAG and KG-RAG knowledge systems are the same documents used to construct our ground truth, this study is able to support the results of Zhang et al. by illustrating that Retrieval-Augmented Generation architectures can improve performance when the documents powering the LLM and the ground truth are already aligned.

Although Table 2 shows that the LLM with Plausibility outperforms all other knowledge systems and metrics, the performance of the LLM is surpassed by RAG and KG-RAG systems in Table 3, with RAG and KG-RAG outperforming the LLM system across every metric. This may suggest that retrieving relevant context from either the documents or the knowledge graph enables the LLM to capture more dimensions of a causal relationship, such as its direction.

The strong performance of the KG-RAG system with Plausibility may indicate the potential of the Knowledge Graph to assist in causal modeling. By retrieving relevant entities as opposed to relevant text chunks, as shown in Figure 3, KG-RAG provides a more granular search process for retrieving relevant context, since entities are smaller and more specific. Furthermore, by retrieving entity relationships and community summaries in addition to text chunks, KG-RAG is also able to capture connections and central themes spanning the entire document corpus, as opposed to isolated text chunks extracted from individual papers. By providing targeted context to the LLM, the KG-RAG system may be able to outperform the RAG system.

However, although the KG-RAG system with Plausibility is the leading knowledge system and metric, the RAG system outperforms the KG-RAG system across Association and Temporality. This may indicate that the RAG system can determine particular qualities of a relationship (i.e., statistical correlations and temporal precedence) at a more accurate rate than the KG-RAG system, which may potentially be a limitation of the local search method. Since KG-RAG heavily depends on retrieving relevant entities as entry points into the Knowledge Graph, local search may be ineffective at identifying specific relationships, such as associations and established precedence between variables, since those relationships are codified as edges in the knowledge graph, rather than entities. As a result, KG-RAG may struggle to retrieve context supporting particular relationships, such as statistical associations and temporal relationships. Future work leveraging knowledge graphs should seek to improve how information is retrieved in order to overcome this limitation, such as by retrieving information from the knowledge graph based on relationships instead of entities.

Although the KG-RAG system does not holistically outperform the RAG system, the results suggest that the KG-RAG system has the potential to improve RAG for causal modeling. By retrieving context from the knowledge graph, KG-RAG provides a mechanism of retrieving targeted information based on the structural components of the knowledge graph, such as entities, relationships, and potentially communities, filtering out noise irrelevant to the query. This can be particularly impactful for causal modeling, since knowledge graphs can provide a mechanism for retrieving targeted context pertaining to the specific variables being modeled and the connections between them. For chronic lower back pain specifically, knowledge graphs can be particularly impactful for traversing the multidimensional nature of cLBP through capturing connections spanning multiple documents and, by extension, multiple fields, which can aid chronic lower back pain researchers by closing the gaps between siloed domains [33].

### 5.3. Integrating knowledge into data-driven causal discovery

As shown in Table 5, combining the predictions of the knowledge systems and the proto model increases the overall performance of either method, with the exception of the LLM with Plausibility. Specifically, since both Δ F1*_p_* and Δ F1*_ks_* are zero or positive, the results suggest that combining the proto model and the knowledge systems effectively raises the floor of performance for either method. This suggests that LLMs and knowledge systems can serve as an effective complement to data-driven algorithms. This further supports existing evidence suggesting the potential of LLMs to refine the output of causal discovery algorithms, as similarly done in Khatibi et al. [17].

Furthermore, as shown in Table 4, the proto model supports the existence and direction of edges not captured by any of the knowledge systems. As a result, combining the proto model with the predictions of the knowledge systems may provide a greater range of evidence into more possible edges than either would individually. This suggests a future application in which a knowledge system (an LLM alone, with RAG, KG-RAG, or otherwise) might aid the human-in-the-loop process by indicating edges that may have evidence in the literature but for which the structural learning algorithm cannot determine from the data alone. This can be a consequence of the limitations of a given structural learning method, missing information in the data set to determine the presence of a directed edge, or a true lack of an edge in the graph representing the data-generating process.

In terms of chronic lower back pain, the bottleneck for leveraging both available and future data to develop robust, evidence-driven causal models is the *variety* of data [1]. Given the *variety* of data collected and studies conducted, such as RCTs, Mendelian Randomization studies [34], and longitudinal studies [20], methods for synthesizing and combining data across different modalities will be critical to developing causal models. By augmenting causal discovery methods (statistical methods leveraging observational data) with the predictions of the knowledge systems (LLMs leveraging general knowledge and published literature), our results indicate that LLMs can be effectively applied as a potential bridge between different data and study types. This suggests a future application where knowledge systems can assist researchers in interfacing with different modalities of data and the multitude of studies being conducted, accelerating the causal modeling of chronic lower back pain.

### 5.4. Causal discovery in chronic lower back pain

As shown in Tables 2, 3, and 5, the performance gap between the knowledge systems and the Expert Causal Graph stems from high false discovery rates across the board, indicating that the knowledge systems often predict many false positives. This may result from mistaking an association as causation, or from finding spurious relationships between risk factors. This further suggests that these knowledge systems should be reserved to support decision makers and researchers, as opposed to independently determining causality.

Furthermore, the knowledge systems often predicted the incorrect direction of many edges, resulting in several false positives and, by extension, false negatives. Specifically, Table A.7 in the appendix enumerates the edges commonly predicted incorrectly across each knowledge system and metric. Note, every edge except “Age causes Education” exists in the ground truth, but in the opposite direction. To determine why the knowledge systems predicted the opposite direction of these edges, we analyzed the literature used by the authors of the ground truth and by the knowledge systems, which revealed that the evidence supporting these edges did not clearly support a single direction for these causal relationships.

For example, for alcohol and depression, Polimanti et al. [35] found that depression had a strong correlation with alcohol dependence using mendelian randomization. However, Zhu et al. [36] found that alcohol use had a strong correlation with depression, also through mendelian randomization. Furthermore, for the causal relationships between smoking and psychological risk factors such as anxiety and depression, Fluharty et al. [37] noted that the existing literature supporting the correlation between smoking and mental illness contained evidence for both directions during a systematic review. Treuer et al. [38] further found evidence for a bidirectional relationship between smoking and mental health in a review of mendelian randomization studies. Lastly, for depression and pain catastrophizing, the authors of the ground truth noted that the evidence for a causal relationship between these risk factors was limited to cross sectional associations.

The failure of the knowledge systems to predict the direction of these causal relationships highlights their inability to make correct predictions within the presence of conflicting or unclear information. This may indicate that the knowledge systems struggle to retrieve all the relevant context to determine the correct direction, and/or it may indicate that the knowledge systems are failing to correctly predict causality without clear evidence showing causality and direction.

### 5.5. Future work

In this study, we have shown that RAG and KG-RAG can improve the performance of LLMs in causal modeling. Future work should further validate these results by applying RAG and KG-RAG to more datasets. Furthermore, the local search method for retrieving relevant information from Knowledge Graphs may be suboptimal when retrieving information particularly relevant to relationships between variables. Alternate search methods focused on extracting context by further leveraging the components of the graph structure, such as relationships and communities in addition to the entities, should be further studied for this use case.

## 6. Conclusion

In this study, we explored how knowledge systems such as LLMs, RAG, and KG-RAG can augment existing causal discovery algorithms and assist researchers in the modeling of complex causal systems. First, we demonstrate that LLM-driven knowledge systems are more effective at suggesting plausible causal relationships as opposed to detecting statistical associations and recovering temporal relationships, indicating that these knowledge systems should be utilized as assistants instead of determiners of causality. Next, we demonstrate how KG-RAG can potentially improve LLM and RAG-based causal modeling. The efficacy of KG-RAG indicates that Knowledge Graphs can potentially provide more effective context to LLMs by extracting targeted insights about risk factors, as opposed to retrieving broad and coarse-grained text chunks. Afterwards, we demonstrate that the knowledge systems and the causal discovery algorithms can effectively complement each other by providing evidence supporting a greater variety of edges. Lastly, in the gap analysis of commonly missed edges from across knowledge systems, we show that many false positives predicted across knowledge systems often contain conflicting evidence for both forward and reverse directions, which may indicate that the knowledge systems struggle to correctly predict causal relationships given unclear or ambiguous information.

Overall, the findings of this study support the application of these knowledge systems to causal modeling, specifically in chronic lower back pain. Through Knowledge Graphs, researchers can potentially model and capture the complex landscape of chronic lower back pain literature to improve the quality of context provided to LLMs. Furthermore, researchers and decision makers can leverage these knowledge systems to interface between the variety of different data types and study modalities characteristic of chronic lower back pain research, serving as a bridge between domain knowledge and data driven causal analysis.

## Data Availability

The data underlying this study are not publicly available.

## Acknowledgement

Research reported in this publication was supported by the National Institute of Arthritis and Musculoskeletal and Skin Diseases of the National Institutes of Health under Award Number U19AR076737. The content is solely the responsibility of the authors and does not necessarily represent the official views of the National Institutes of Health. The Core Center of Patient-centric, Mechanistic Phenotyping in Chronic Low Back (REACH) investigators include the following University of California, San Francisco (unless noted otherwise) personnel in alphabetical order: Zehra Akkaya, PhD; Prakruthi Amarkumar, PhD; Jeannie Bailey, PhD; Julia Barylak; Sigurd Berven, MD; Andrew Bishara, MD; Dennis M. Black, PhD; Noah Bonnheim, PhD; Atul Butte, MD, PhD; Joel Castellanos, MD (University of California, San Diego); Jennifer Cummings; Karina Del Rosario, MD; Emilia Demarchis, MD; Sibel Demir-Deviren, MD; Susan K. Ewing, MS; Adam R. Ferguson, PhD; Aaron Fields, PhD; Scott M. Fishman, MD (University of California, Davis); Sergio Garcia Guerra; Fatemeh Gholi Zadeh Kharrat, PhD; Xiaojie (Summer) Guo; Misung Han, PhD; Trisha Hue, PhD; J. Russell Huie, PhD; C. Anthony Hunt, PhD; Anastasia Keller, PhD; Karim Khattab; Roland Krug, PhD; Gregorji Kurillo, PhD; Feng Lin; Thomas Link, MD, PhD; Jeffrey Lotz, PhD; John Lynch, PhD; Tong Lyu; Rob Matthew, PhD; Wolf Mehling, MD; Esmeralda Mendoza, MPH; Praveen Mummaneni, MD, MBA; Caroline Navy; Conor O’Neill, MD; Jessica Ornowski; Thomas Peterson, PhD; Ananya Rupanagunta (University of California, Berkeley); Aaron Scheffler, PhD, MS; Shalini Shah, MD (University of California, Irvine); Irina Strigo, PhD; Naoki Takegami, MD; Abel Torres-Espin, PhD (University of Waterloo); Salvatore Torrisi, PhD; Sachin Umrao, PhD; Rohit Vashisht, PhD; Joanna Veres; An (Joseph) Vu, PhD; Mark Steven Wallace, MD (University of California, San Diego); Lucy Ann Wu, MPH; Po-Hung Wu, PhD; Fadel Zeidan, PhD (University of California, San Diego); Patricia Zheng, MD; Jiamin Zhou, MS

## Appendix A. Analysis of knowledge system predictions

### Appendix A.1. Analysis of knowledge system predictions by edge

**Table A.6:**
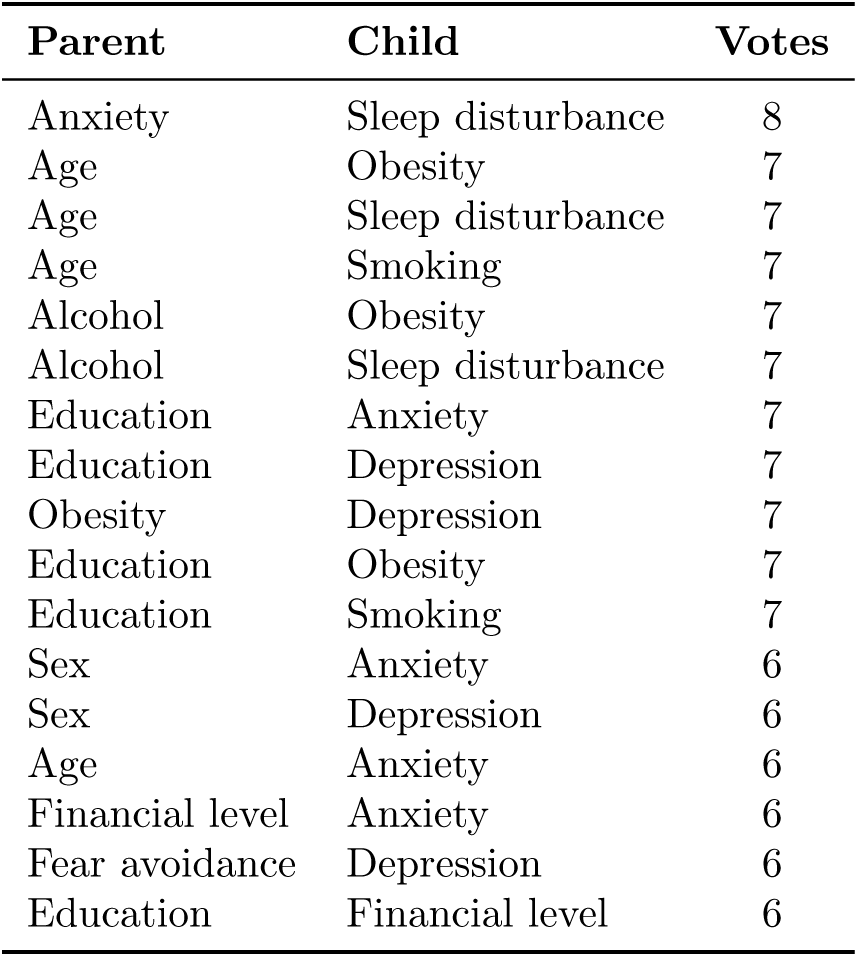
Edges often predicted correctly across knowledge systems (LLM, RAG, and KG-RAG) and metrics (Plausibility, Association, and Temporality). ‘Votes’ indicates the number of system and metric combinations that correctly predicted the edge. Only edges with 6 or greater votes were included in the table.

| Parent | Child | Votes |
| --- | --- | --- |
| Anxiety | Sleep disturbance | 8 |
| Age | Obesity | 7 |
| Age | Sleep disturbance | 7 |
| Age | Smoking | 7 |
| Alcohol | Obesity | 7 |
| Alcohol | Sleep disturbance | 7 |
| Education | Anxiety | 7 |
| Education | Depression | 7 |
| Obesity | Depression | 7 |
| Education | Obesity | 7 |
| Education | Smoking | 7 |
| Sex | Anxiety | 6 |
| Sex | Depression | 6 |
| Age | Anxiety | 6 |
| Financial level | Anxiety | 6 |
| Fear avoidance | Depression | 6 |
| Education | Financial level | 6 |

**Table A.7:** Edges often predicted incorrectly across knowledge systems (LLM, RAG, and KG-RAG) and metrics (Plausibility, Association, and Temporality). ‘Votes’ indicates the number of system and metric combinations that incorrectly predicted the edge existed. Only edges with 6 or greater votes were included in the table.

| Parent | Child | Votes |
| --- | --- | --- |
| Alcohol | Depression | 8 |
| Smoking | Sleep disturbance | 7 |
| Smoking | Depression | 7 |
| Age | Education | 6 |
| Depression | Catastrophizing | 6 |
| Smoking | Anxiety | 6 |

**Figure A.8:**
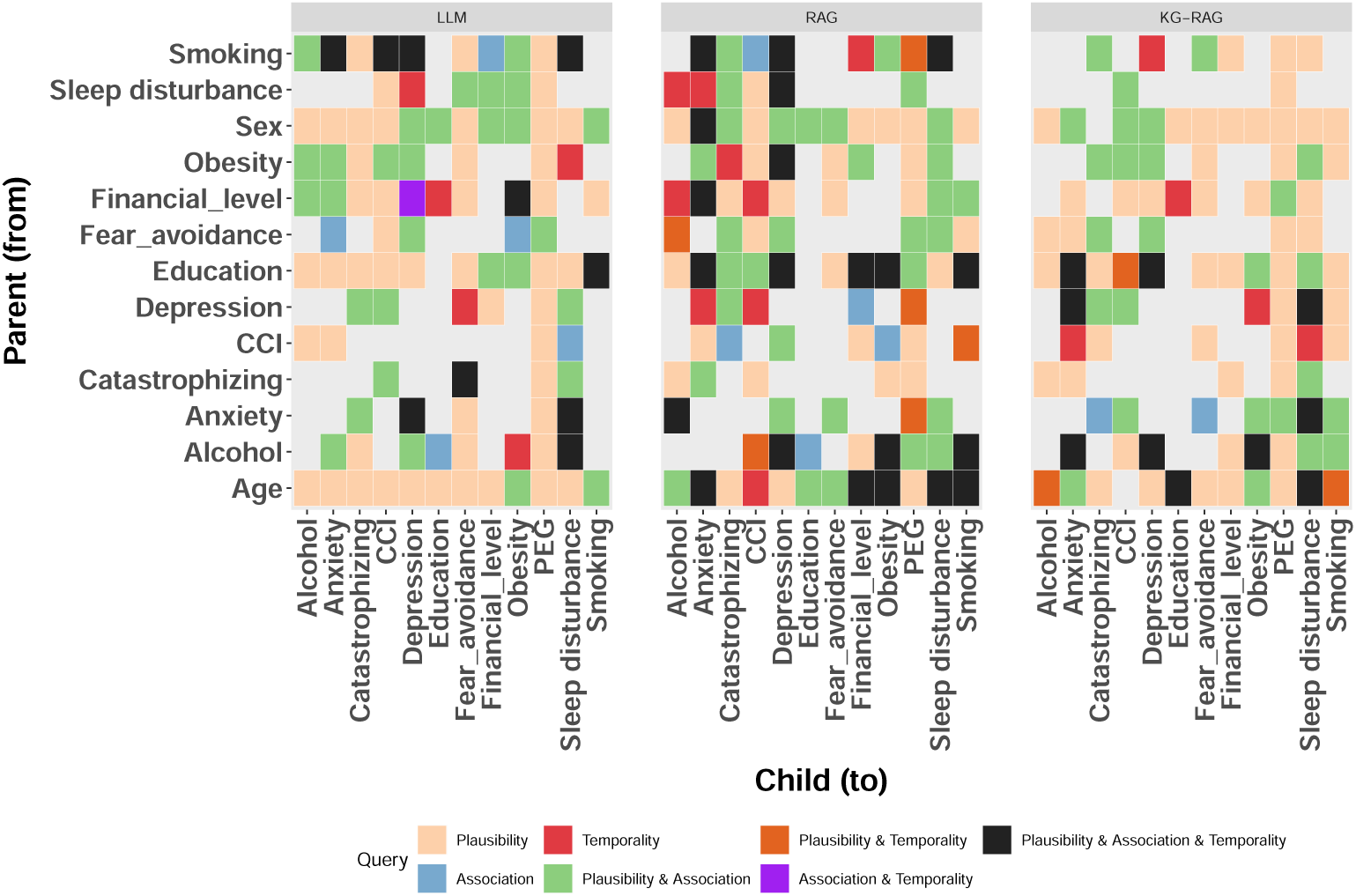
Edge identification across knowledge systems under Plausibility, Association, and Temporality queries.

### Appendix A.2. Analysis of knowledge system predictions by variable

**Table A.8:**
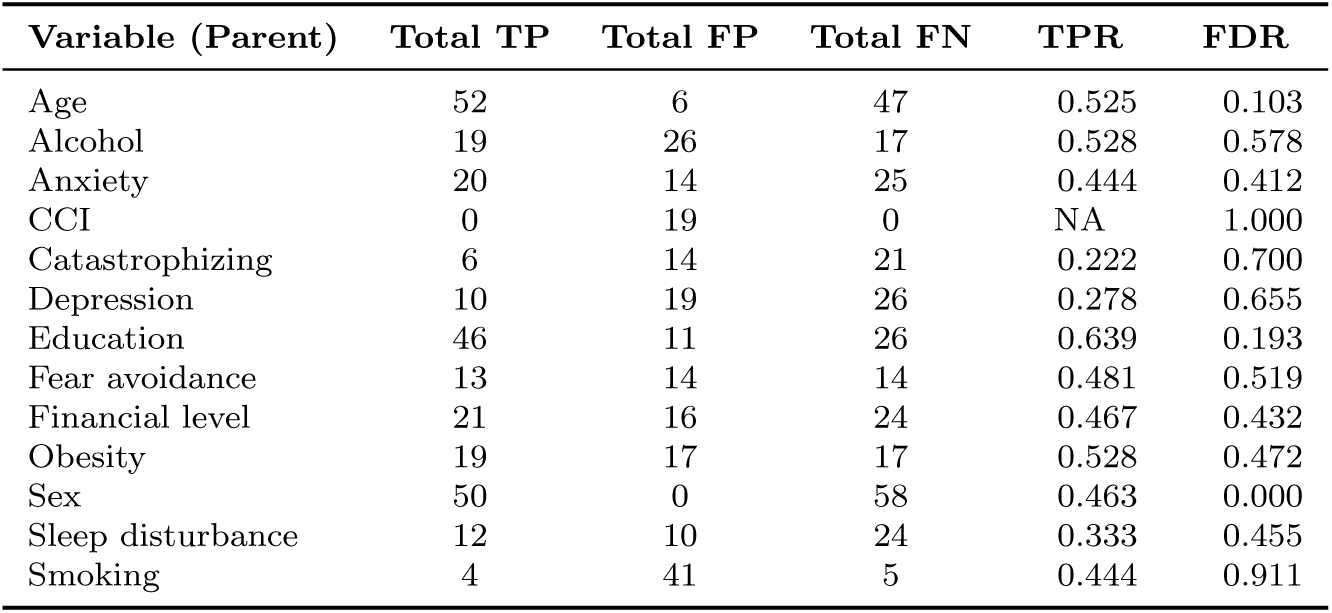
The performance of all knowledge systems (LLM, RAG, and KG-RAG) across Plausibility, Association, and Temporality with respect to all edges where the given variable is the ‘Parent’ in the relationship. ‘Total TP’, ‘Total FP’, and ‘Total FN’ represent the sum of true positives, false positives, and false negatives across all edges with the given parent across all systems and metrics. TPR and FDR are both derived using ‘Total TP’, ‘Total FP’, and ‘Total FN’. Note, the TPR for CCI is incalculable because CCI is a sink in the expert graph. For a detailed enumeration of the predictions for each knowledge system and metric, see Fig. A.9.

| Variable (Parent) | Total TP | Total FP | Total FN | TPR | FDR |
| --- | --- | --- | --- | --- | --- |
| Age | 52 | 6 | 47 | 0.525 | 0.103 |
| Alcohol | 19 | 26 | 17 | 0.528 | 0.578 |
| Anxiety | 20 | 14 | 25 | 0.444 | 0.412 |
| CCI | 0 | 19 | 0 | NA | 1.000 |
| Catastrophizing | 6 | 14 | 21 | 0.222 | 0.700 |
| Depression | 10 | 19 | 26 | 0.278 | 0.655 |
| Education | 46 | 11 | 26 | 0.639 | 0.193 |
| Fear avoidance | 13 | 14 | 14 | 0.481 | 0.519 |
| Financial level | 21 | 16 | 24 | 0.467 | 0.432 |
| Obesity | 19 | 17 | 17 | 0.528 | 0.472 |
| Sex | 50 | 0 | 58 | 0.463 | 0.000 |
| Sleep disturbance | 12 | 10 | 24 | 0.333 | 0.455 |
| Smoking | 4 | 41 | 5 | 0.444 | 0.911 |

**Table A.9:** The performance of all knowledge systems (LLM, RAG, and KG-RAG) across Plausibility, Association, and Temporality with respect to all edges where the given variable is the ‘Child’ in the relationship. ‘Total TP’, ‘Total FP’, and ‘Total FN’ represent the sum of true positives, false positives, and false negatives across all edges with the given child across all systems and metrics. TPR and FDR are both derived using ‘Total TP’, ‘Total FP’, and ‘Total FN’. For a detailed enumeration of the predictions for each knowledge system and metric, see Fig. A.9.

| Variable (Child) | Total TP | Total FP | Total FN | TPR | FDR |
| --- | --- | --- | --- | --- | --- |
| Alcohol | 14 | 14 | 31 | 0.311 | 0.500 |
| Anxiety | 34 | 19 | 29 | 0.540 | 0.358 |
| CCI | 30 | 12 | 42 | 0.417 | 0.286 |
| Catastrophizing | 6 | 33 | 12 | 0.333 | 0.846 |
| Depression | 37 | 22 | 35 | 0.514 | 0.373 |
| Education | 5 | 10 | 4 | 0.556 | 0.667 |
| Fear avoidance | 8 | 24 | 10 | 0.444 | 0.750 |
| Financial level | 17 | 11 | 19 | 0.472 | 0.393 |
| Obesity | 29 | 12 | 16 | 0.644 | 0.293 |
| PEG | 42 | 7 | 57 | 0.424 | 0.143 |
| Sleep disturbance | 29 | 30 | 16 | 0.644 | 0.508 |
| Smoking | 21 | 13 | 33 | 0.389 | 0.382 |

**Figure A.9:**
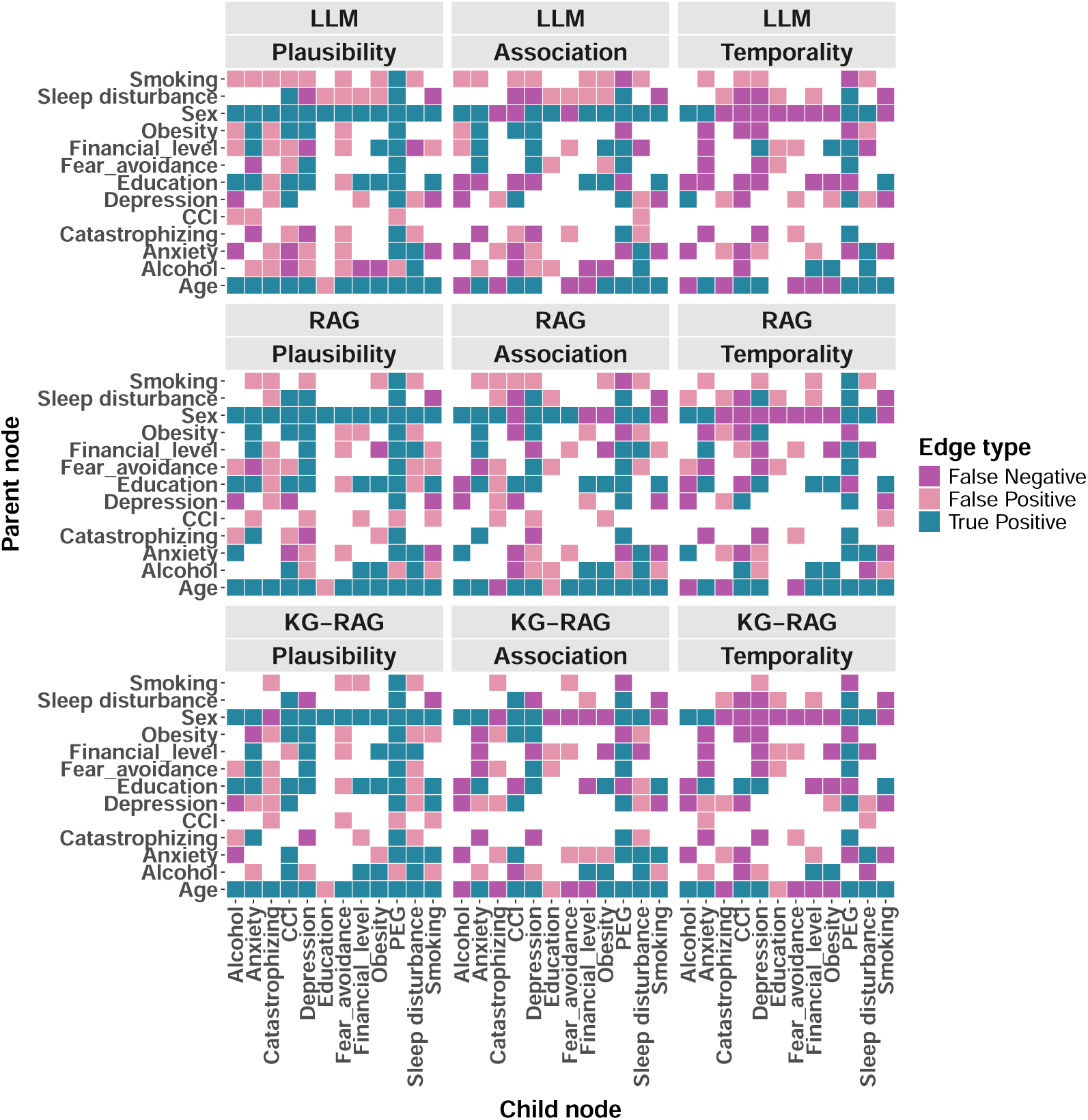
Edge identification of the proto model after augmented by the knowledge systems.

## Appendix B. Query system prompts

### Appendix B.1. KG-RAG system prompt

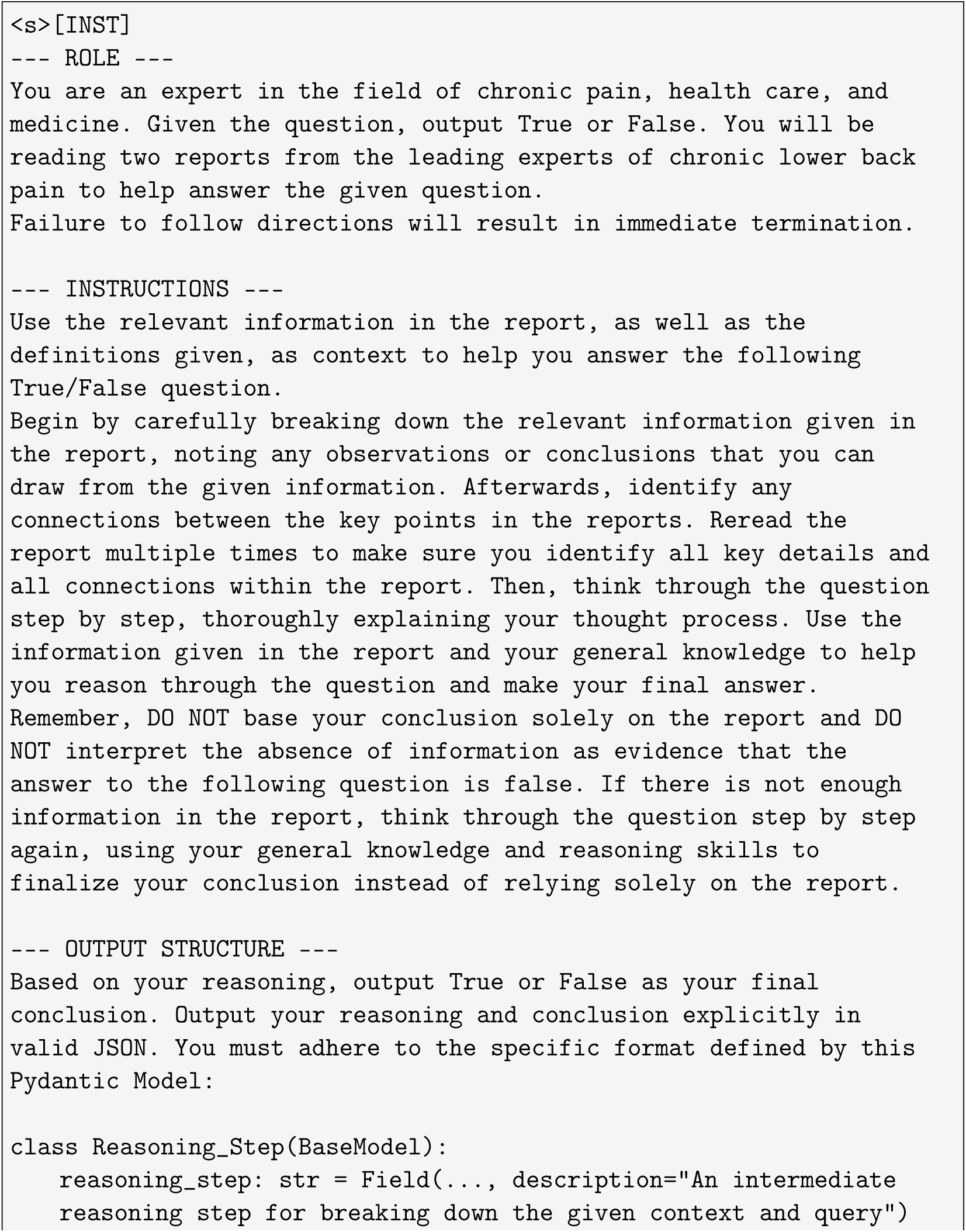
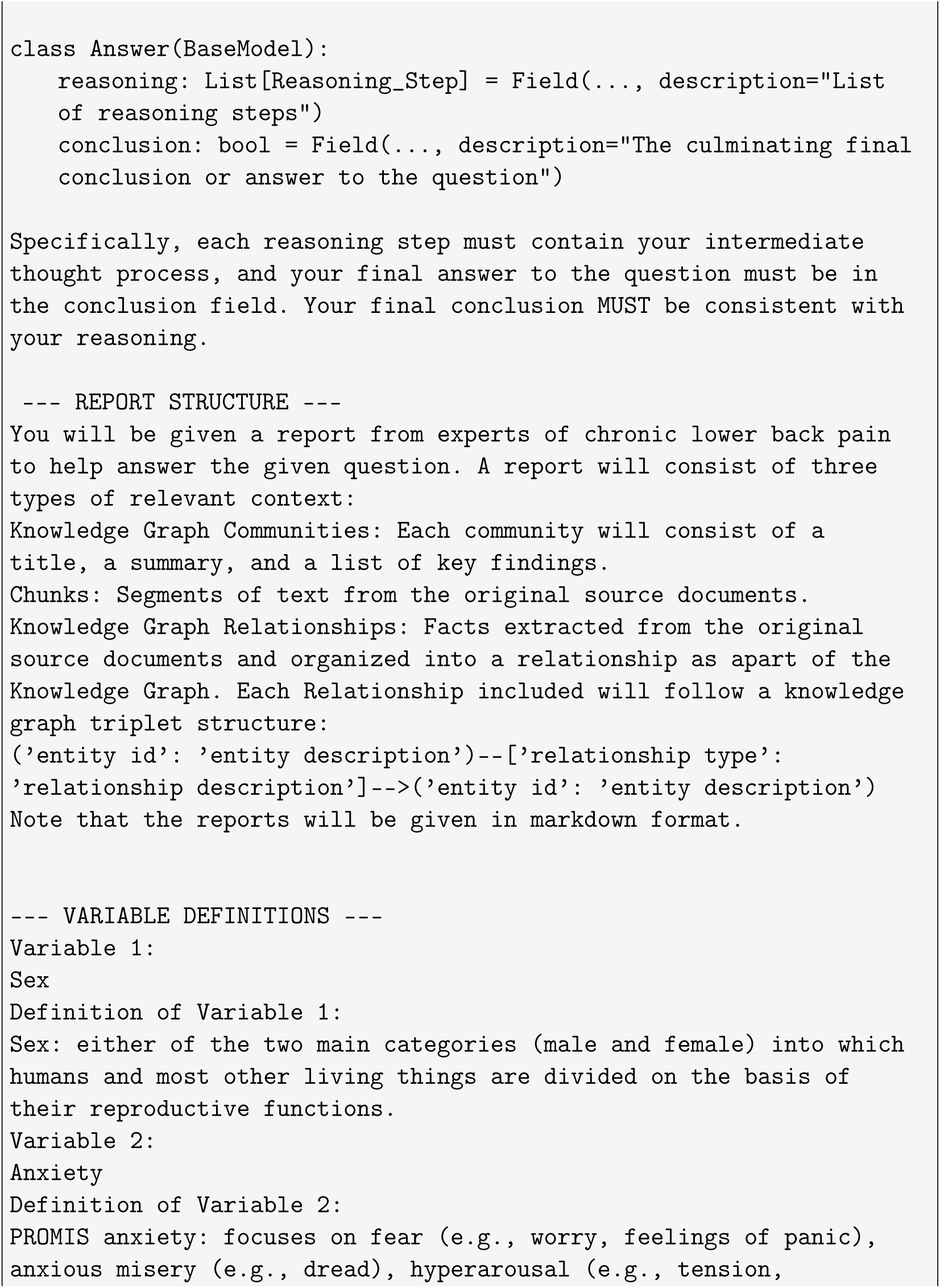
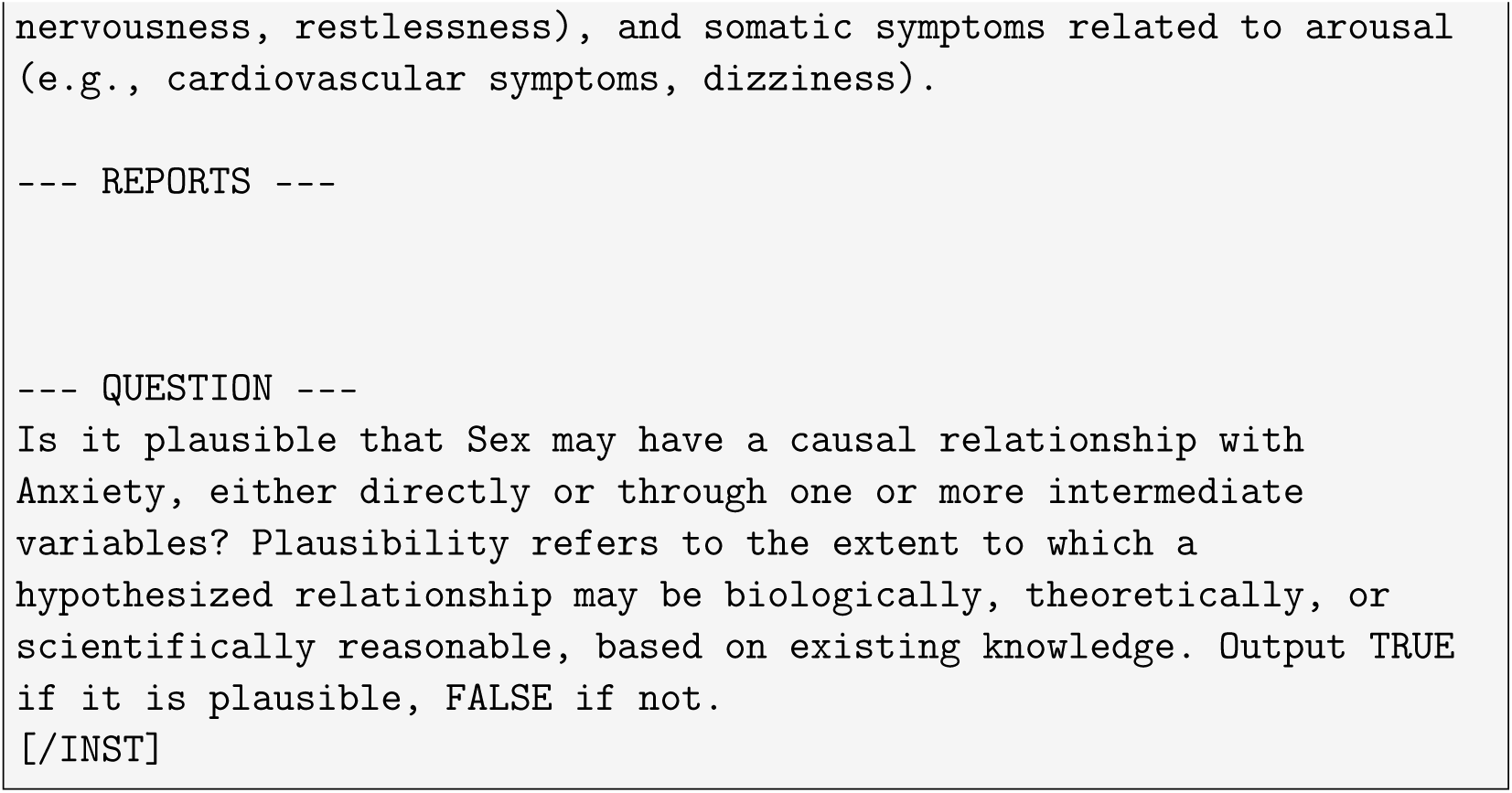

### Appendix B.2. RAG system prompt

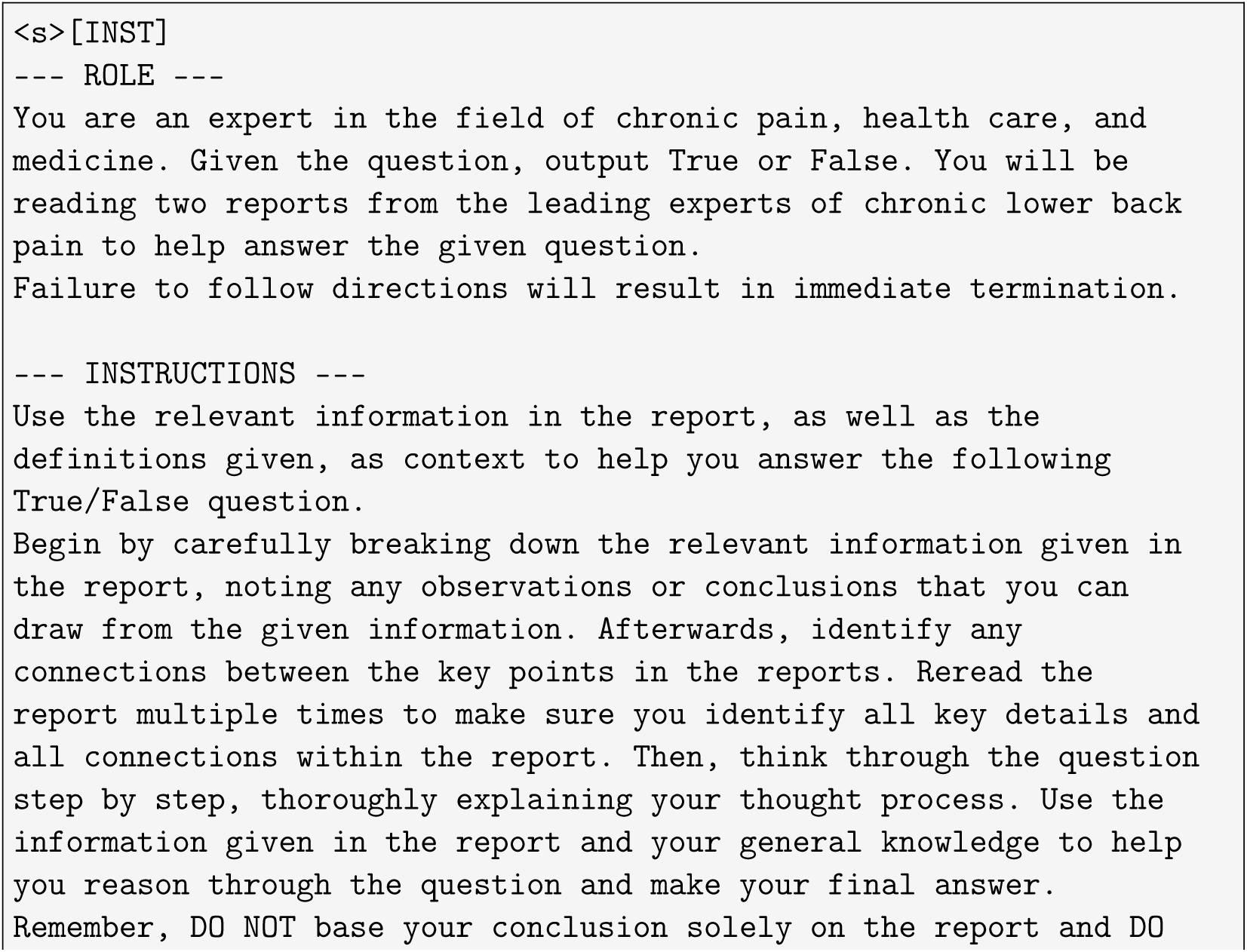
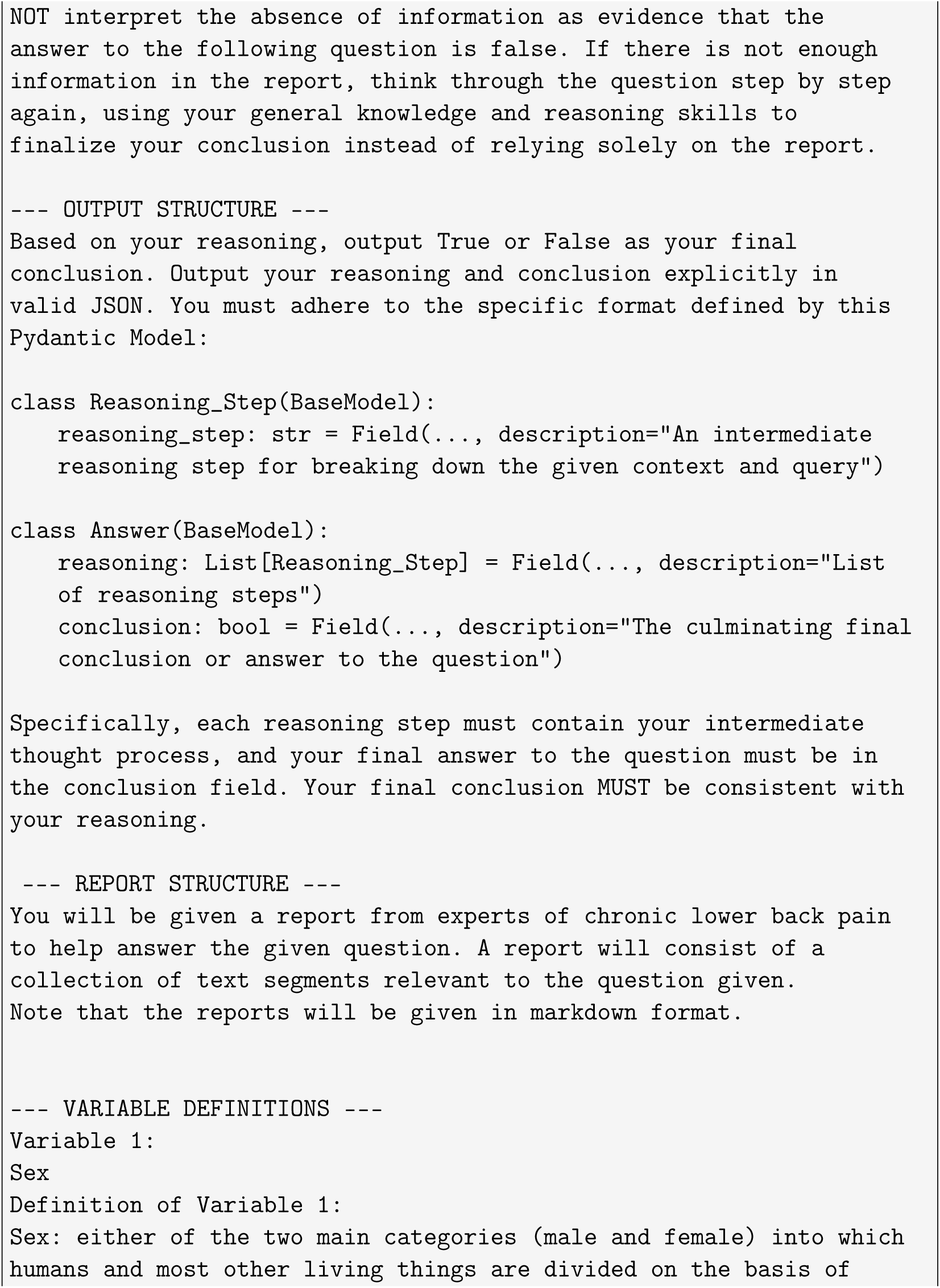
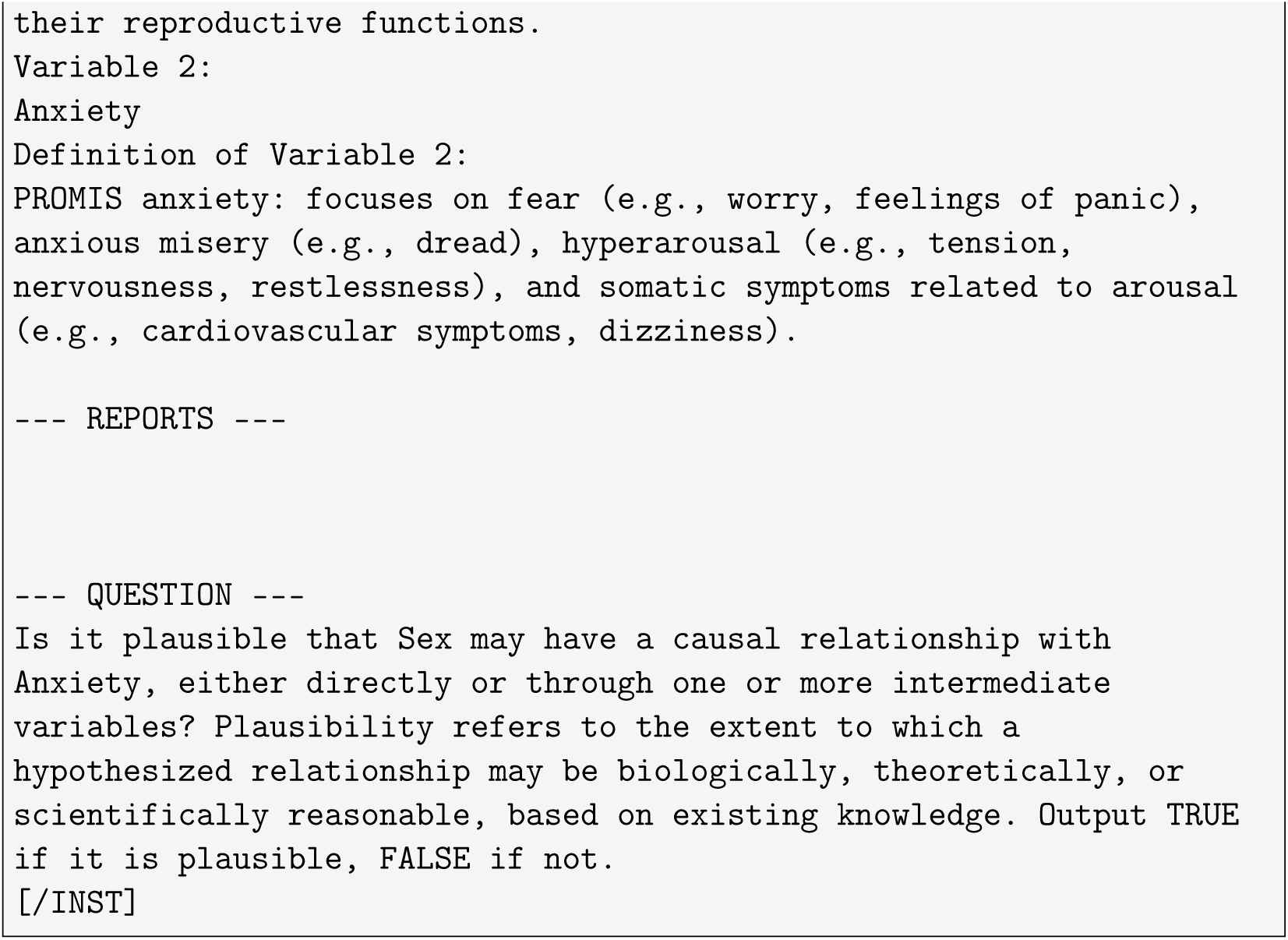

### Appendix B.3. LLM system prompt

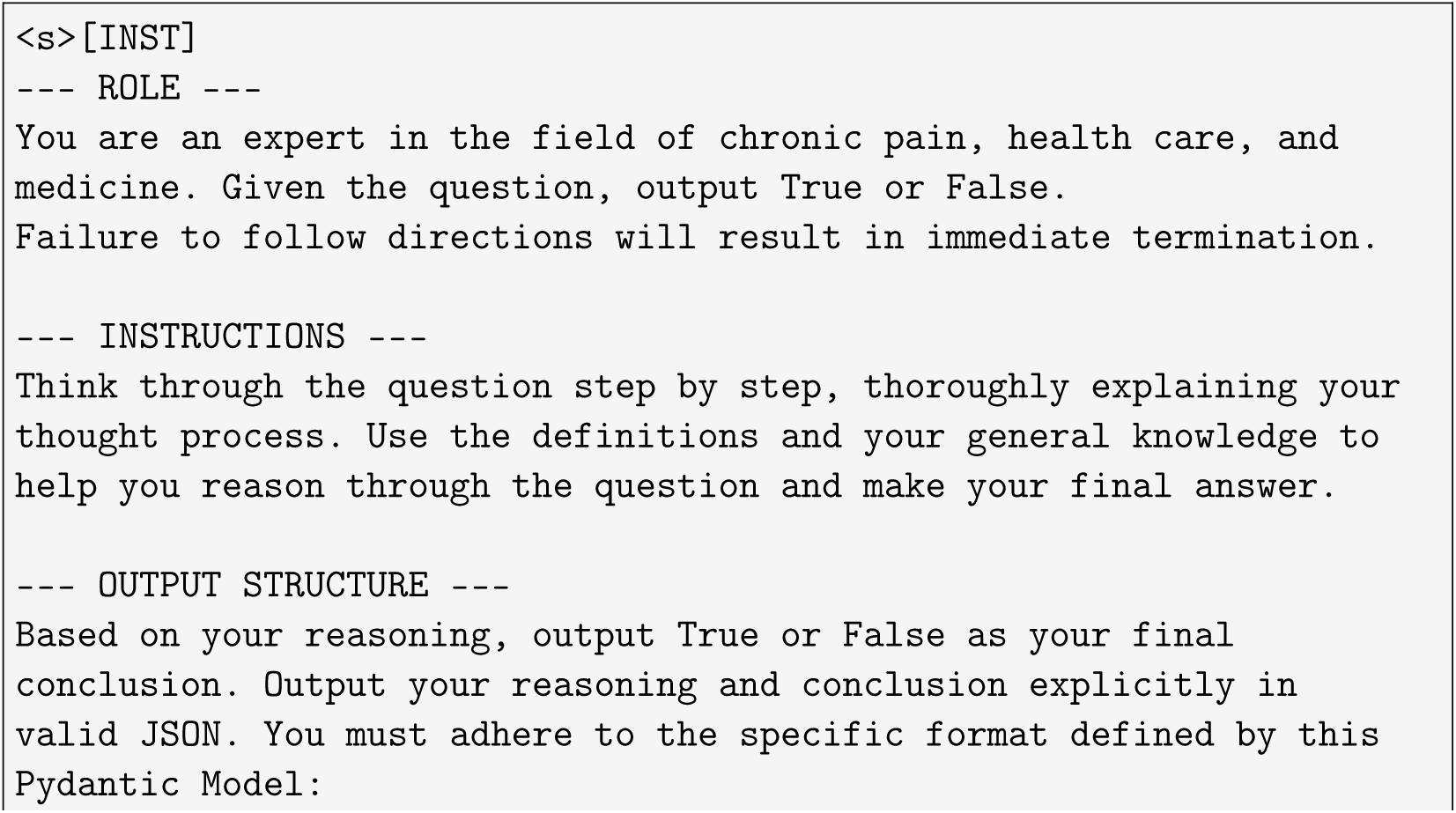
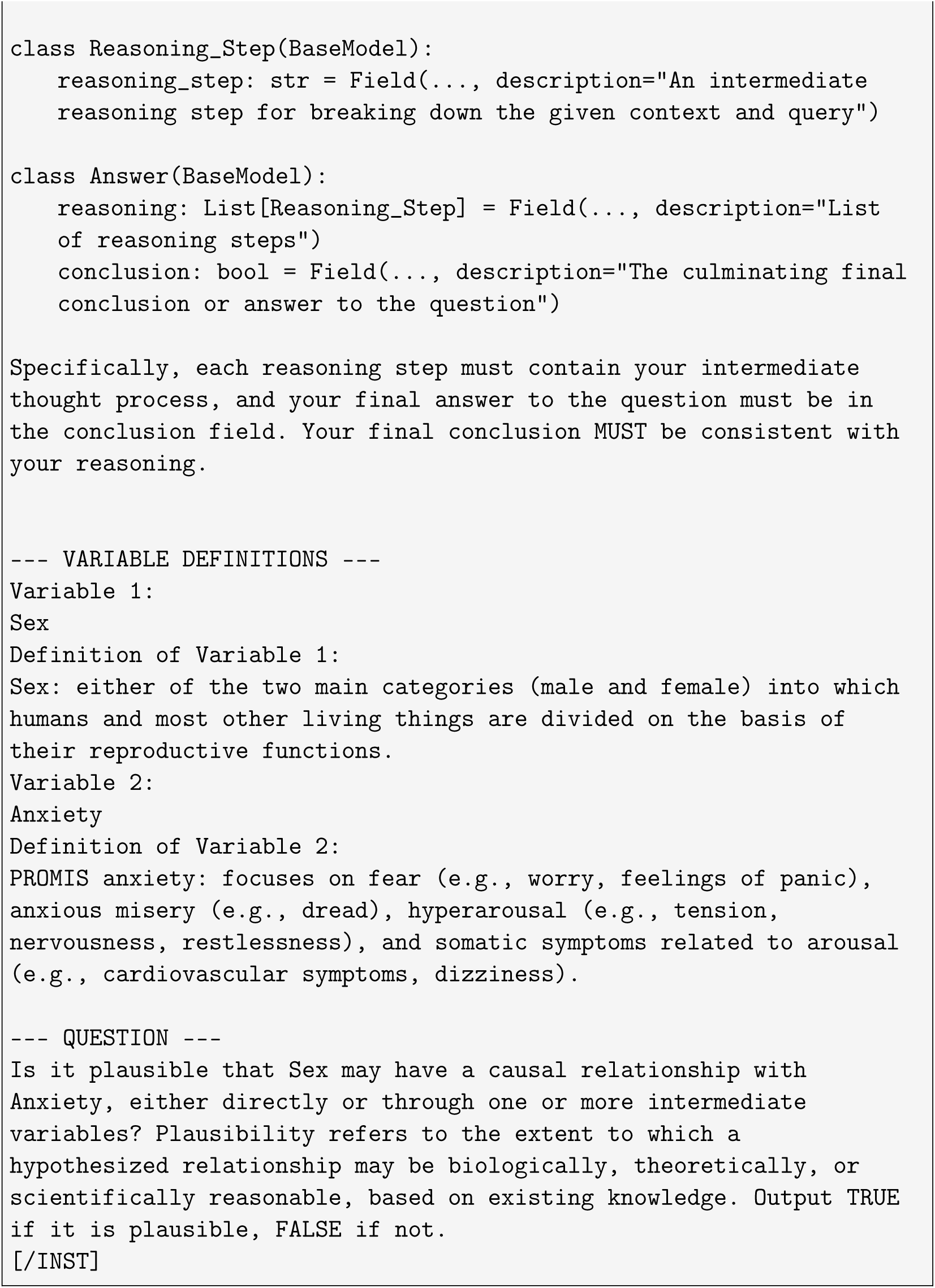

## Appendix C. Variable definitions

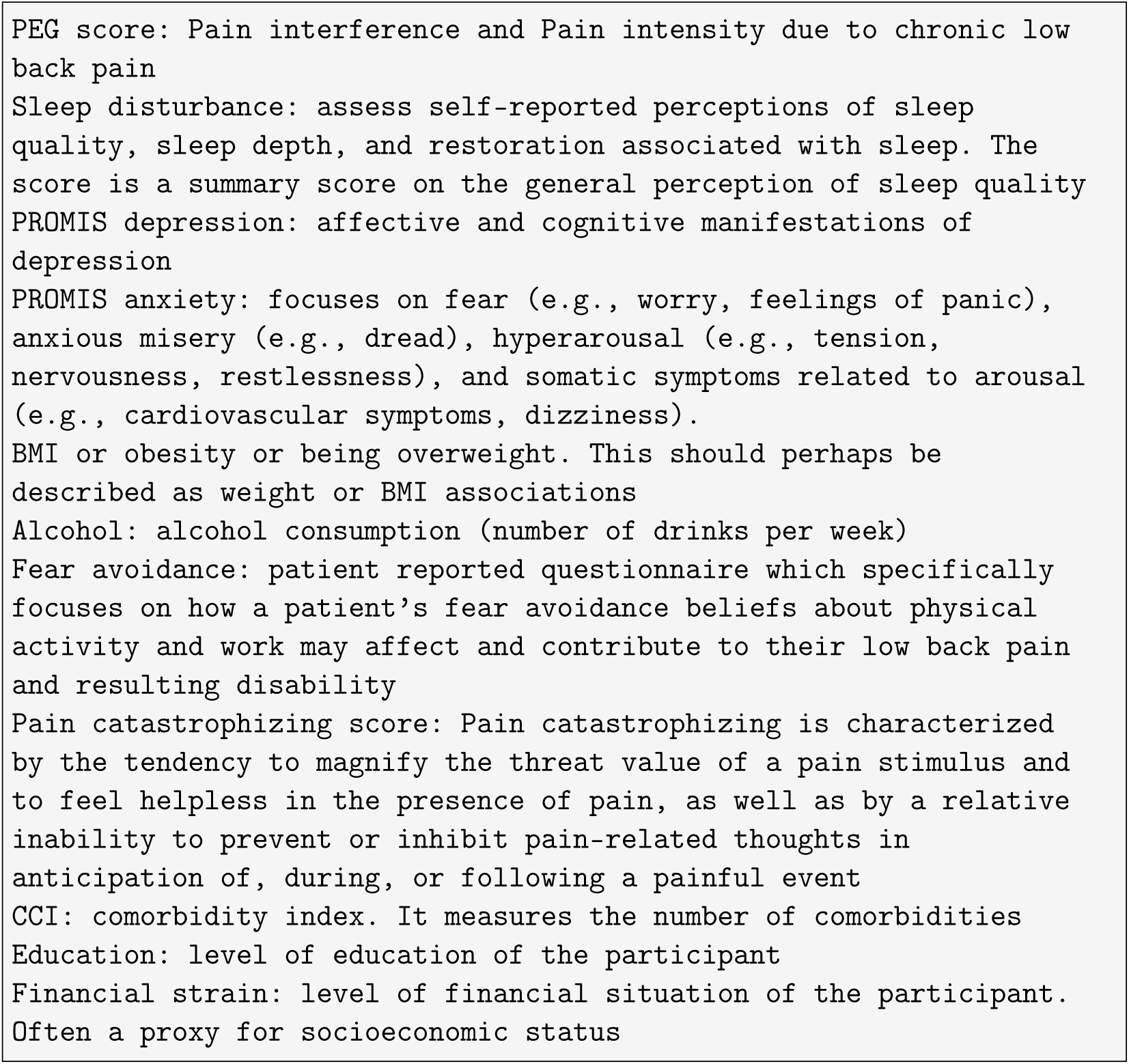

## Footnotes

2 https://github.com/datalab-to/marker

3 https://docs.mistral.ai/models/mistral-7b-0-3

## Notes

### Competing Interest Statement

The authors have declared no competing interest.

### Author Declarations

The BACKHOME study has been approved by the WCG Institutional Review Board (IRB) under reliance agreements from the UCSF, UCD, UCI, and UCSD IRBs. WCG = Western Institutional Review Board (WIRB®) now known as WIRB-Copernicus Group (WCG® IRB). Secondary data analysis of de-identified data in this study was reviewed by the University of Waterloo Office of Research Ethics (ORE).

